# Nationwide large-scale data of acute lower gastrointestinal bleeding in Japan uncover detailed etiologies and relevant outcomes: CODE BLUE J-Study

**DOI:** 10.1101/2021.01.18.21250035

**Authors:** Naoyoshi Nagata, Katsumasa Kobayashi, Atsushi Yamauchi, Atsuo Yamada, Jun Omori, Takashi Ikeya, Taiki Aoyama, Naoyuki Tominaga, Yoshinori Sato, Takaaki Kishino, Naoki Ishii, Tsunaki Sawada, Masaki Murata, Akinari Takao, Kazuhiro Mizukami, Ken Kinjo, Shunji Fujimori, Takahiro Uotani, Minoru Fujita, Hiroki Sato, Sho Suzuki, Toshiaki Narasaka, Junnosuke Hayasaka, Tomohiro Funabiki, Yuzuru Kinjo, Akira Mizuki, Shu Kiyotoki, Tatsuya Mikami, Ryosuke Gushima, Hiroyuki Fujii, Yuta Fuyuno, Naohiko Gunji, Yosuke Toya, Kazuyuki Narimatsu, Noriaki Manabe, Koji Nagaike, Tetsu Kinjo, Yorinobu Sumida, Sadahiro Funakoshi, Kana Kawagishi, Tamotsu Matsuhashi, Yuga Komaki, Kuniko Miki, Kazuhiro Watanabe, Masakatsu Fukuzawa, Takao Itoi, Naomi Uemura, Takashi Kawai, Mitsuru Kaise

**Author notes:** **Correspondence:** Naoyoshi Nagata, MD, PhD., Department of Gastroenterological Endoscopy, Tokyo Medical University, 6-7-1, Nishishinjuku, Shinjuku-ku, Tokyo 160-0023, Japan.

## Abstract

**Background:** The value of endoscopy for acute lower GI bleeding (ALGIB) remains unclear, given few large cohort studies. We aim to provide detailed clinical data for ALGIB management and to identify patients at risk for adverse outcomes based on endoscopic diagnosis.

**Methods:** We conducted a multicenter, retrospective cohort study, named CODE BLUE J-Study, in 49 hospitals throughout Japan and studied 10,342 cases admitted for outpatient-onset of acute hematochezia.

**Results:** Cases were mostly elderly, with 29.5% hemodynamic instability and 60.1% comorbidity. 69.1% and 87.7 % of cases underwent CT and colonoscopy, respectively. Diagnostic yield of colonoscopy reached 94.9%, revealing 48 etiologies, most frequently diverticular bleeding. During hospitalization, the endoscopic therapy rate was 32.7%, mostly using clipping and band ligation. IVR and surgery were infrequently performed, for 2.1% and 1.4%. In-hospital rebleeding and death occurred in 15.2% and 0.9%. Diverticular bleeding cases had higher rates of hemodynamic instability, rebleeding, endoscopic therapy, IVR, and transfusion, but lower rates of death and surgery than other etiologies. Small bowel bleeding cases had significantly higher rates of surgery, IVR, and transfusion than other etiologies. Malignancy or upper GIB cases had significantly higher rates of thromboembolism and death than other etiologies. Etiologies that have favorable outcomes were ischemic colitis, infectious colitis, and post-endoscopy bleeding.

**Conclusions:** Large-scale data of patients with acute hematochezia revealed high proportions of colonoscopy and CT, resulting in high endoscopic therapy rates. We highlight the importance of colonoscopy in detecting accurate bleeding etiologies that stratify patients at high or low risk of adverse outcomes.

## Introduction

Acute lower gastrointestinal bleeding (ALGIB) manifests as relatively mild hematochezia (maroon or red blood passed through the rectum) progressing to massive hemorrhage with shock[1–3]. Approximately 30-50% of adults with ALGIB will progress to severe bleeding[4–7], and bleeding can recur frequently, requiring re-examination, re-hospitalization, and repeated transfusion[1–7]. Moreover, ALGIB episodes have been shown to increase the risk of subsequent thromboembolism and death, regardless of antithrombotic drug use[8]. Under these conditions, ALGIB presents a significant economic burden[9]. Unlike upper GIB (UGIB), which can be treated by anti-acid therapy, there are no effective therapies for preventing ALGIB and its recurrence[1]. Therefore, approaches to initial management, diagnosis, intervention, and follow-up of ALGIB remain uncertain, and there is likely to be considerable variation in clinical practice. Although the rate of UGIB has been decreasing rapidly over the past 10 years, LGIB has increased slightly[10,11], which also applies to Japan[12]. Therefore, evidence-building for the management of ALGIB will grow increasingly important.

Although a few guidelines for ALGIB exist[1–3], most of statements are supported by low to moderate quality evidence because of challenges in performing randomized controlled trials (RCTs) in the ALGIB setting[13]. Even if a RCT can be performed, it may differ from clinical practice because elderly patients with comorbid conditions are frequently excluded from RCTs[14,15]. We therefore consider it important to clarify the factors associated with outcomes from observational studies without excluding patients. However, an adequate sample size is required to analyze the outcome and its associated factors because small sample size may result in insufficient power and type 2 error[16,17]. To date, compared to UGIB, there are few large studies using real-world clinical data, especially more than 1,000 ALGIB cases. Although one large UK study showed detailed clinical courses in ALGIB[18], it could not accurately identify bleeding etiologies by endoscopy, or assess their relevant outcomes. Adverse clinical outcomes may substantially differ among etiologies; if so, this confirms the value of colonoscopy, as recommended in the guidelines[1,3].

To address these issues, we have conducted a multi-center observational cohort study throughout Japan, collecting data from over 10,000 cases of acute hematochezia and extensively examining ALGIB etiologies. We aim to provide detailed clinical data for ALGIB management, to identify patients at risk for adverse outcomes by bleeding etiology, and to compare clinical data to the UK study.

## Methods

### Study design, settings, and participants

We conducted a multicenter, retrospective cohort study in emergency hospitals throughout Japan. To collect real-world clinical data, we invited the participation of gastroenterology physicians who were directly involved in the treatment of hematochezia. A total of 49 facilities located in 25 prefectures, from Okinawa prefecture in the south to Aomori prefecture in the north, wanted to participate. Representative physicians in each hospital agreed to a study of detailed clinical data of patients with acute hematochezia, named CODE BLUE-J Study (COlonic DivErticular Bleeding Leaders Update Evidence from multicenter Japanese Study). The ethics committees and the institutional review boards approved conducting this study with the opt-out method in all 49 participating hospitals (**Supplementary Table 1**). Patients and/or the public were not involved in the design, or conduct, or reporting, or dissemination plans of this research.

**Table 1.** Baseline characteristics.

| **Factor** | **Value** | **Data available for analysis** | **Missing value** |
| --- | --- | --- | --- |
| Outpatient onset | 10,342 (100) | 10,342 | 0 |
| Number of ambulances | 5,859 (3,854-9,246) | 49 | 0 |
| 24/7 colonoscopy access | 49 (100) | 49 | 0 |
| Age | 74 (63-82) | 10,342 | 0 |
| Age ≥ 60 y | 8,327 (80.5) | 10,342 | 0 |
| Male | 6,317 (61.1) | 10,342 | 0 |
| Blood type, O | 3,312 (33.7) | 9,841 | 501 (4.8) |
| Blood type, A | 3,644 (37.0) | 9,841 | 501 (4.8) |
| Blood type, B | 2,021 (20.5) | 9,841 | 501 (4.8) |
| Blood type, AB | 864 (8.8) | 9,841 | 501 (4.8) |
| Height, cm | 160 (152-167) | 9,789 | 553 (5.3) |
| Body weight, kg | 57.3 (48.9-66.4) | 9,921 | 421 (4.1) |
| BMI | 22.5 (20.1-24.9) | 9,715 | 627 (6.1) |
| BMI > 25 | 2,381 (24.5) | 9,715 | 627 (6.1) |
| Alcohol, current drinker | 4,130 (46.3) | 8,918 | 1424 (13.8) |
| Smoking, never | 4,702 (51.2) | 9,179 | 1163 (11.2) |
| Smoking, current | 1,661 (18.1) | 9,179 | 1163 (11.2) |
| Smoking, ever | 2,816 (30.7) | 9,179 | 1163 (11.2) |
| Performance status, 1 | 8,976 (87.8) | 10,220 | 122 (1.2) |
| Performance status, 2 | 694 (6.8) | 10,220 | 122 (1.2) |
| Performance status, 3 | 304 (3.0) | 10,220 | 122 (1.2) |
| Performance status, 4 | 244 (2.4) | 10,220 | 122 (1.2) |
| Blood pressure (mmHg) | 127 (111-145) | 10,161 | 181 (1.8) |
| Heart rate (/minutes) | 83 (73-96) | 10,140 | 202 (2) |
| Syncope/ loss of consciousness | 668 (6.5) | 10,324 | 18 (0.2) |
| Hemodynamic instability* | 3,046 (29.5) | 10,342 | 0 |
| Abdominal pain | 1,664 (16.1) | 10,323 | 19 (0.2) |
| Fever | 660 (6.4) | 10,320 | 22 (0.2) |
| Diarrhea | 1,016 (9.9) | 10,307 | 35 (0.3) |
| Hematochezia | 10,342 (100) | 10,342 | 0 |
| Tarry stools | 593 (5.8) | 10,321 | 21 (0.2) |
| Hemoglobin (g/dl) | 11.4 (9.3-13.1) | 10,334 | 8 (0.1) |
| Hemoglobin ≤ 7.0 (g/dl) | 798 (7.7) | 10,342 | 0 |
| White blood cells (/ul) | 7,150 (5,600-9,300) | 10,335 | 7 (0.1) |
| Platelet count (/ul) | 20.8 (16.8-25.3) | 10,331 | 11 (0.1) |
| Albumin (g/dl) | 3.7 (3.3-4.1) | 9,857 | 485 (4.7) |
| PT-INR | 1.0 (1.0-1.0) | 9,012 | 1330 (12.9) |
| Hematocrit (%) | 34.2 (28.4-39) | 10,322 | 20 (0.2) |
| Blood urea nitrogen (mg/dl) | 19.0 (14.7-25.0) | 10,273 | 69 (0.7) |
| Creatinine (mg/dl) | 0.8 (0.7-1.1) | 10,269 | 73 (0.7) |
| CRP (mg/dl) | 0.2 (0.1-0.5) | 10,067 | 275 (2.7) |
| History of bowel resection | 752 (7.3) | 10,340 | 2 (0.01) |
| History of chemotherapy | 338 (3.3) | 10,319 | 23 (0.2) |
| History of radiation therapy | 241 (2.3) | 10,325 | 17 (0.2) |
| Past history LGIB | 3,090 (30.0) | 10,342 | 0 |
| History of angiectasia | 72 (0.7) | 10,341 | 1 (0.01) |
| History of IBD | 233 (2.3) | 10,341 | 1 (0.01) |
| History of diverticular bleeding | 2,603 (25.2) | 10,334 | 8 (0.1) |
| History of ischemic colitis | 223 (2.2) | 10,341 | 1 (0.01) |
| Charlson Comorbidity Index, 0 | 4,124 (39.9) | 10,342 | 0 |
| Charlson Comorbidity Index, 1 | 2,431 (23.5) | 10,342 | 0 |
| Charlson Comorbidity Index, $\geq 2$ | 3,787 (36.6) | 10,342 | 0 |
| Diabetes mellitus, uncomplicated | 1,933 (18.7) | 10,342 | 0 |
| Diabetes mellitus, end-organ damage | 350 (3.4) | 10,342 | 0 |
| Hemiplegia | 278 (2.7) | 10,334 | 8 (0.1) |
| Cerebral vascular accident or TIA | 1,475 (14.3) | 10,340 | 2 (0.02) |
| COPD | 315 (3.1) | 10,342 | 0 |
| Dementia | 565 (5.5) | 10,334 | 8 (0.1) |
| Connective tissue disease | 418 (4.0) | 10,342 | 0 |
| Myocardial infarction | 1,660 (16.1) | 10,342 | 0 |
| Chronic heart failure | 854 (8.3) | 10,340 | 2 (0.02) |
| Peptic ulcer disease | 726 (7.0) | 10,342 | 0 |
| Moderate chronic kidney disease | 1,479 (14.3) | 10,340 | 2 (0.02) |
| Severe chronic kidney disease | 326 (3.2) | 10,342 | 0 |
| Peripheral vascular disease | 421 (4.1) | 10,342 | 0 |
| Leukemia/ myeloma | 68 (0.7) | 10,342 | 0 |
| AIDS | 19 (0.2) | 10,333 | 9 (0.1) |
| Solid tumor, localized | 1,332 (12.9) | 10,335 | 7 (0.1) |
| Solid tumor, metastatic | 254 (2.5) | 10,342 | 0 |
| Liver disease, mild | 217 (2.1) | 10,341 | 1 (0.01) |
| Liver disease, moderate to severe | 207 (2.0) | 10,341 | 1 (0.01) |
| Malignant lymphoma | 83 (0.8) | 10,342 | 0 |
| Hypertension | 5,842 (56.5) | 10,342 | 0 |
| Dyslipidemia | 2,822 (27.3) | 10,341 | 1 (0.01) |
| NSAIDs | 1,177 (11.4) | 10,342 | 0 |
| NSAIDs (noncoxib) | 929 (9.0) | 10,342 | 0 |
| COX-2 selective inhibitors | 272 (2.6) | 10,342 | 0 |
| Low-dose aspirin | 2,056 (20.0) | 10,342 | 0 |
| Thienopyridine | 1,014 (9.8) | 10,342 | 0 |
| Cilostazol | 243 (2.4) | 10,342 | 0 |
| Other antiplatelet drugs | 303 (2.9) | 10,342 | 0 |
| Number of antiplatelet drugs, 0 | 7,420 (71.8) | 10,342 | 0 |
| Number of antiplatelet drugs, 1 | 2,262 (21.9) | 10,342 | 0 |
| Number of antiplatelet drugs, 2 | 626 (6.1) | 10,342 | 0 |
| Number of antiplatelet drugs, 3 | 34 (0.3) | 10,342 | 0 |
| Warfarin | 705 (6.8) | 10,342 | 0 |
| DOACs | 615 (6.0) | 10,342 | 0 |
| Acetaminophen | 277 (2.7) | 10,342 | 0 |
| Corticosteroid | 579 (5.6) | 10,342 | 0 |

Note. \*Hemodynamic instability was defined as initial systolic blood pressure < 90 mm Hg, initial heart rate ≥ 100/min, or the presence of syncope.
Abbreviations. IQR, interquartile range; BMI, body mass index; PT-INR, international normalized ratio of prothrombin time; CRP, C-reactive protein; LGIB, lower gastrointestinal bleeding; IBD, inflammatory bowel disease; TIA, transient ischemic attack; COPD, chronic obstructive pulmonary disease; AIDS, acquired immune deficiency syndrome; NSAIDs, nonsteroid anti-inflammatory drugs; COX, Celecoxib; DOACs, direct oral anticoagulants.

We selected patients admitted for acute hematochezia from January 2010 to December 2019. We included patients aged ≥20 years at the onset of hematochezia regardless of presence of tarry stools, diarrhea, abdominal pain, or fever. We excluded patients after elimination of hematochezia, those who were hospitalized for examination for anemia, whose onset of hematochezia was in hospital, or for whom data collection was not possible due to insufficient information on medical records. A total of 11,035 patients were included, and all data were re-evaluated rigorously by secretariat’s institution (Tokyo Medical University); then 693 patients met the exclusion criteria. Ultimately, 10,342 cases emergently admitted for outpatient onset of acute, continuous, or frequent hematochezia were evaluated.

### Data collection

Before collecting data, we conducted research meetings three times with representatives from the 49 participating hospitals on the content and definition of survey items. At the research meetings, it was agreed to aim for the registration of at least 100 cases from each institution. The survey items were prepared using Excel sheets formatted to define each clinical factor and data entry rule, and sent to each participating institution. To prevent data omissions or entry errors, and to reduce missing values, we used the data-validation rules in Excel to input the values and unknowns for the categorical variables (e,g, diabetes mellitus, 0, 1, unknown), and free input for the continuous variables (**Supplementary Table 2**). At each hospital, data were collected from electric endoscopic records and medical records, and input into an Excel sheet. The sheet was sent to the secretariat’s facility (Tokyo Medical University). This facility evaluated omissions and errors (e.g., admission or endoscopy date, data error value) in the input values for the data sent from each hospital (**Supplementary Table 2**). The data to be corrected were communicated by the input physician(s) of each institution with detailed comments. Such communication was conducted more than three times per hospital with the use of Excel sheets by e-mail.

**Table 2.** CT and endoscopic diagnosis.

| **Factor** | **Value** | **Data available for analysis** | **Missing value** |
| --- | --- | --- | --- |
| CT of the abdomen/pelvis | 7,149 (69.1) | 10,342 | 0 |
| Contrast enhanced CT | 5,240 (73.3) | 7,149 | 0 |
| Urgent CT | 6,968 (97.5) | 7,149 | 0 |
| Time to CT scan, hours | 1 (1-2) | 7,149 | 0 |
| Extravasation on CT | 1,151 (22.0) | 5,240 | 0 |
| Extravasation at jejunum | 7 (0.1) | 5,240 | 0 |
| Extravasation at ileum | 57 (1.1) | 5,240 | 0 |
| Extravasation at cecum | 53 (1.0) | 5,240 | 0 |
| Extravasation at ascending | 547 (10.4) | 5,240 | 0 |
| Extravasation at transverse | 83 (1.6) | 5,240 | 0 |
| Extravasation at descending | 138 (2.6) | 5,240 | 0 |
| Extravasation at sigmoid | 224 (4.3) | 5,240 | 0 |
| Extravasation at rectum | 49 (0.9) | 5,240 | 0 |
| Extravasation at upper GI tract | 1 (0.02) | 5,240 | 0 |
| Colonic diverticular bleeding on CT | 1,846 (25.8) | 7,149 | 0 |
| Enterocolitis on CT | 953 (13.3) | 7,149 | 0 |
| Tumor lesion on CT | 90 (1.3) | 7,149 | 0 |
| Other diagnosis on CT | 190 (2.7) | 7,149 | 0 |
| Initial colonoscopic examination | 9,066 (87.7) | 10,342 | 0 |
| Time to 1<sup>st</sup> colonoscopy, hours | 16 (4-32) | 9,066 | 0 |
| SRH on initial endoscopy | 2,801 (30.9) | 9,066 | 0 |
| SRH, active bleeding | 1,489 (16.4) | 9,066 | 0 |
| SRH, visible vessel | 535 (5.9) | 9,066 | 0 |
| SRH, adherent clot | 829 (9.1) | 9,066 | 0 |
| Location of SRH, cecum | 163 (1.8) | 9,066 | 0 |
| Location of SRH, ascending | 1,166 (12.9) | 9,066 | 0 |
| Location of SRH, transverse | 262 (2.9) | 9,066 | 0 |
| Location of SRH, descending | 182 (2.0) | 9,066 | 0 |
| Location of SRH, sigmoid | 630 (7.0) | 9,066 | 0 |
| Location of SRH, rectum | 349 (3.9) | 9,066 | 0 |
| Location of SRH, jejunum | 5 (0.06) | 9,066 | 0 |
| Location of SRH, ileum | 94 (1.0) | 9,066 | 0 |
| Location of SRH, upper GI | 1 (0.01) | 9,066 | 0 |
| Secondary colonoscopic examination | 1,992 (19.2) | 10,342 | 0 |
| Time to 2<sup>nd</sup> colonoscopy, hours | 53 (29-96) | 1,992 | 0 |
| **Final diagnosis** |  |  |  |
| Colonic diverticular bleeding | 6,575 (63.6) | 10,342 | 0 |
| Definitive diverticular bleeding | 2,386 (23.1) | 10,342 | 0 |
| Presumptive diverticular bleeding | 4,189 (40.5) | 10,342 | 0 |
| Ischemic colitis | 941 (9.1) | 10,342 | 0 |
| Post-procedure bleeding | 463 (4.5) | 10,342 | 0 |
| Post-ESD bleeding | 140 (1.4) | 10,342 | 0 |
| Post-polypectomy bleeding | 73 (0.7) | 10,342 | 0 |
| Post-EMR bleeding, | 223 (2.2) | 10,342 | 0 |
| Post-biopsy bleeding | 16 (0.2) | 10,342 | 0 |
| Post-other procedure bleeding | 12 (0.1) | 10,342 | 0 |
| Rectal ulcer | 257 (2.5) | 10,342 | 0 |
| IBD | 210 (2.0) | 10,342 | 0 |
| Hemorrhoid bleeding | 184 (1.8) | 10,342 | 0 |
| Colorectal angioectasia | 133 (1.3) | 10,342 | 0 |
| Colorectal malignancy | 193 (1.9) | 10,342 | 0 |
| Colorectal cancer | 168 (1.6) | 10,342 | 0 |
| Metastatic tumor | 16 (0.2) | 10,342 | 0 |
| Other colorectal tumor* | 9 (0.1) | 10,342 | 0 |
| Colorectal polyp | 37 (0.4) | 10,342 | 0 |
| Infectious colitis | 134 (1.3) | 10,342 | 0 |
| Radiation colitis | 66 (0.6) | 10,342 | 0 |
| Non-specific colitis | 47 (0.5) | 10,342 | 0 |
| Drug-induced ulcer | 14 (0.1) | 10,342 | 0 |
| Non-specific ulcer | 56 (0.5) | 10,342 | 0 |
| Colorectal varix | 25 (0.2) | 10,342 | 0 |
| Dieulafoy ulcer | 12 (0.1) | 10,342 | 0 |
| Postoperative anastomotic bleeding | 14 (0.1) | 10,342 | 0 |
| Anal bleeding other than hemorrhoids** | 12 (0.1) | 10,342 | 0 |
| Diverticulitis | 7 (0.1) | 10,342 | 0 |
| Small bowel bleeding | 246 (2.4) | 10,342 | 0 |
| Small bowel bleeding (definitive) | 114 (1.1) | 10,342 | 0 |
| Small bowel bleeding (presumptive) | 121 (1.2) | 10,342 | 0 |
| Meckel's diverticular bleeding | 11 (0.1) | 10,342 | 0 |
| UGIB | 153 (1.5) | 10,342 | 0 |
| Other diagnosis*** | 37 (0.4) | 10,342 | 0 |
| Unknown | 526 (5.1) | 10,342 | 0 |
| Missing data | 0 | 10,342 | 0 |

Note. \*Other tumor included 4 diseases; malignant lymphoma (n=2), gastrointestinal stromal tumor (n=1), pseudomyxoma of the appendix (n=1), and submucosal tumor unknown origin (n=5).
\*\*Anal bleeding other than hemorrhoids included 3 diseases; anal laceration or fissure (n=10), post-anal surgery bleeding (n=1), and anal condyloma (n=1).
\*\*\*Other diagnosis included 15 diseases; mucosal bleeding (n=8), mucosal prolapse syndrome (n=6), colorectal laceration (n=4), fistula or penetration into colorectum (n=3), colorectal perforation (n=2), mucosal lymphoid hyperplasia (n=2), Kaposi's sarcoma (n=1), stoma-related bleeding (n=2), pseudoaneurysm (n=2), intussusception (n=1), postoperative stenosis (n=1), graft-versus-host disease (n=1), hematoma (n=2), Henoch-Schönlein purpura (n=1), Cronkhite-Canada syndrome (n=1).
Abbreviations. IQR, interquartile range; CT, computer tomography; SRH, stigmata of recent hemorrhage; ESD, endoscopic submucosal dissection; EMR, endoscopic mucosal resection;
IBD, inflammatory bowel disease; UGIB, upper gastrointestinal bleeding.

### Variables and outcomes

Detailed clinical data on patient characteristics and management of hematochezia during hospitalization and after discharge were assessed. As for clinical data, 219 survey items were considered essential (**Supplementary Table 2**), and 49 items were optional. Baseline characteristics consisted of 75 items, including presenting symptoms, vital signs, blood sample data, past histories, comorbidities, and drug intake within 30 days of admission (**Supplementary Table 2**). We evaluated 19 comorbidities with the Charlson comorbidity index (CCI)[19], which has been validated and commonly used for GIB research[6,8,18]. Moreover, comorbidities of hypertension and dyslipidemia, not included in CCI, were also collected. During hospitalization, information of CT and endoscopic diagnosis consisting of 80 items (e.g., stigmata of recent hemorrhage (SRH) on endoscopy, etiology of bleeding) was collected. We also evaluated procedures consisting of 41 items, such as type of endoscopic therapy, IVR, and surgery (**Supplementary Table 2**). Because patients may have received two different or identical procedures during hospitalization due to events after examination and treatment, we evaluated for the procedure items twice. Final diagnosis was made mainly by initial and second endoscopy. Small bowel endoscopy, capsule endoscopy, and upper GI endoscopy were performed at the discretion of each institution based on symptoms and tests. Clinical outcomes, consisting of 23 items, included rebleeding, thromboembolism, and mortality (**Supplementary Table 2**). Dates of occurrence of outcomes were evaluated during hospitalization and after discharge. After discharge, we followed up patients from the index date to the occurrence of any clinical outcomes, and censored patients at the time of the last visit, end of follow-up, or death. In the survival analysis, the endpoint was death, and data were censored as of the time of the last visit, or the end of follow-up. Rebleeding and second rebleeding were evaluated and defined as significant amounts of fresh, bloody or wine-colored stools after admission[6,8,18]. We defined a thromboembolic event as the development of cardiovascular events, cerebrovascular events, pulmonary embolism, or deep vein thrombosis[8]. The diagnosis of thromboembolism was based on typical symptoms, with imaging modalities including CT, magnetic resonance imaging, coronary angiography, ventilation-perfusion scan, ultrasonography, or electrocardiography[8]. Date and cause of death were ascertained from death certificates and medical record reviews[8]. Cause of death was additionally determined from laboratory tests, multiple imaging modalities, or autopsy[8]. Secondary outcomes were blood transfusion during hospital stay and prolonged length of hospital stay (LOS).

### Statistical methods

Descriptive statistics, reported as number and percentage, or median and inter quartile range (IQR) are used to describe patient characteristics, procedures, and clinical outcomes. To compare clinical data between two groups, we used the chi-square test or Fisher’s exact test for categorical variables, as appropriate. For the analysis of the association between bleeding etiologies and outcomes, etiologies that have > 50 cases were included. P-values less than 0.05 were considered significant. All statistical analysis was performed using STATA version 14 software (StataCorp, College Station, TX).

## Results

### Baseline characteristics

The median number of ambulances in the 49 participating hospitals was 5,859, and 24/7 colonoscopy was available in all institutions (**Table 1**). The median age was 74 years, with 61.1% male. About half of patients were current drinkers (46.3%) or current or ever smokers (48.8%). Hemodynamic instability rate was 29.5%. All patients presented with hematochezia, with additional existing symptoms of 16.1% abdominal pain, 6.4% fever, and 9.9% diarrhea. Overall, 30% of patients had prior histories of acute LGIB. 60.1% of patients had a CCI ≥1, and the most common comorbidities were hypertension (56.5%), dyslipidemia (27.3%), and diabetes (18.7%). Median laboratory values were 7,150 (/ul) white blood cells, 11.4 (g/dl) hemoglobin, and 3.7 (g/dl) albumin. At presentation, drug administration rates were 11.4% NSAIDs, 20.0% low-dose aspirin, 9.8% thienopyridine, 6.8% warfarin, 6.0% DOACs, 2.7% acetaminophen, and 5.6% corticosteroids. 6.1% of patients took dual antiplatelet therapy.

### CT diagnosis and endoscopic diagnosis

CT of the abdomen/pelvis was undertaken in 69.1% of all cases, with median timing of 1 hour of arriving at the hospital (**Table 2**). Of these, 73.3% undertook contrast enhanced CT, and extravasation findings on CT were identified in 22.0%. Among locations, the ascending colon (10.4%) was most commonly identified in extravasation on CT, more than twice as often as the sigmoid colon (4.3%).

Initial colonoscopy was undertaken in 87.7% of all cases, with median timing of 16 hours of arrival. SRH on endoscopy was identified in 30.9% of cases, with 16.4% exhibiting active bleeding, 5.9% visible vessels, and 9.2% adherent clots. Similar to CT extravasation, the ascending colon (12.9%) was the most frequent site of SRH identification, again almost twice as often as the sigmoid colon (7.0%). A total of 59.2% (6,117/10,342) of patients underwent both endoscopy and CT for further investigation of the source of bleeding, and only 2.4% (244/10,342) of patients who did not undergo colonoscopy had no CT scan. 19.2% of patients underwent a repeat colonoscopy, with median timing of 53 hours of arriving at the hospital, mainly due to rebleeding. Diagnostic yield of initial and second colonoscopy reached 94.9%, and 22 bleeding-source categories, covering 48 diseases, were found (**Table 2**). The most common final diagnosis was colonic diverticular bleeding (63.6%), followed by ischemic colitis (9.1%), post-endoscopic procedure bleeding (4.5%), and rectal ulcers (2.5%). Bleeding sources other than colon, rectum, and anus were also observed, such as small bowel bleeding (2.4%) and upper GIB (1.5%). The rate of each etiology in the final diagnosis differed slightly compared to the initial endoscopic diagnosis (**Supplementary Table 3**); note especially the decreased rate of unknown cases, and increased definite diverticular bleeding.

**Table 3.** Pre-endoscopic procedures, endoscopic procedures, and non-endoscopic procedures.

| Factor | Value | Data available for analysis | Missing value |
| --- | --- | --- | --- |
| Bowel prep, PEG or enema | 7,595 (83.8) | 9,066 | 0 |
| Bowel prep, PEG | 6,022 (66.4) | 9,066 | 0 |
| Bowel prep, enema | 1,731 (19.1) | 9,066 | 0 |
| Endoscopic cap use | 6,537 (72.1) | 9,066 | 0 |
| Endoscopic cap use, long | 1,822 (20.1) | 9,066 | 0 |
| Endoscopic cap use, short | 4,638 (51.2) | 9,066 | 0 |
| Endoscopic cap use, ST hood | 69 (0.8) | 9,066 | 0 |
| Endoscopic cap use, other | 8 (0.1) | 9,066 | 0 |
| Waterjet scope use | 6,971 (77.0) | 9,066 | 0 |
| PEG use in water jet scope | 390 (4.3) | 9,066 | 0 |
| Conservative therapy after endoscopy | 6,092 (67.2) | 9,066 | 0 |
| Endoscopic therapy | 2,784 (30.7) | 9,066 | 0 |
| Clipping | 1759 (63.8) | 2,784 | 0 |
| Indirect clipping | 1,212 (43.5) | 2,784 | 0 |
| Direct clipping | 547 (19.7) | 2,784 | 0 |
| Band ligation | 674 (24.2) | 2,784 | 0 |
| Snare ligation | 109 (3.9) | 2,784 | 0 |
| HSE | 50 (1.8) | 2,784 | 0 |
| OTSC | 0 | 2,784 | 0 |
| Coagulation | 228 (8.2) | 2,784 | 0 |
| Other endoscopic therapy* | 20 (0.7) | 2,784 | 0 |
| Success of endoscopic therapy | 2,663 (95.7) | 2,784 | 0 |
| Failure of endoscopic therapy | 121 (4.4) | 2,784 | 0 |
| Treatment for failure, clipping | 77 (63.6) | 121 | 0 |
| Treatment for failure, band ligation | 8 (6.6) | 121 | 0 |
| Treatment for failure, HSE | 14 (11.6) | 121 | 0 |
| Treatment for failure, OTSC | 1 (0.8) | 121 | 0 |
| Treatment for failure, coagulation | 8 (6.6) | 121 | 0 |
| Treatment for failure, other therapy | 11 (9.1) | 121 | 0 |
| Post-endoscopy perforation | 13 (0.1) | 9,066 | 0 |
| Post-endoscopy diverticulitis | 3 (0.03) | 9,066 | 0 |
| IVR | 143 (1.4) | 10,342 | 0 |
| Surgery | 101 (1.0) | 10,342 | 0 |
| Barium impaction therapy | 66 (0.6) | 10,342 | 0 |
| Endoscopic therapy during hospitalization | 3,379 (32.7) | 10,342 | 0 |
| IVR need during hospitalization | 217 (2.1) | 10,342 | 0 |
| Surgery need during hospitalization | 142 (1.4) | 10,342 | 0 |

Note. Pre-endoscopic and endoscopic procedures were evaluated among patients who underwent endoscopy (n=9,066). \*Other endoscopic therapy included hot biopsy, polypectomy,
endoscopic mucosal resection.
Abbreviations. IQR, interquartile range; PEG, polyethylene glycol; HSE, hypertonic saline epinephrine; OTSC, over the scope clip; IVR, interventional radiology

**Table 4.** Clinical outcomes.

**Tables 4 Clinical outcomes**
| **Factor** | **Value** | **Data available for analysis** | **Missing value** |
| --- | --- | --- | --- |
| In-hospital rebleeding | 1,573 (15.2) | 10,342 | 0 |
| 2nd rebleeding during hospitalization | 458 (4.4) | 10,342 | 0 |
| Out-of-hospital rebleeding | 2,650 (25.6) | 10,342 | 0 |
| Num of out-of-hospital rebleeding episodes, 1-4 | 2,372 (22.9) | 10,342 | 0 |
| Num of out-of-hospital rebleeding episodes, 5-9 | 178 (1.7) | 10,342 | 0 |
| Num of out-of-hospital rebleeding episodes, $\geq 10$ | 100 (1.0) | 10,342 | 0 |
| Occurrence of thromboembolism | 65 (0.6) | 10,342 | 0 |
| Occurrence of acute coronary syndrome | 19 (0.2) | 10,342 | 0 |
| Occurrence of cerebrovascular disease | 28 (0.3) | 10,342 | 0 |
| Occurrence of PE/ DVT | 18 (0.2) | 10,342 | 0 |
| In-hospital death | 97 (0.9) | 10,342 | 0 |
| GI bleeding related death | 13 (13.4) | 97 | 0 |
| Out-of-hospital death | 694 (6.8) | 10,245 | 0 |
| Follow-up days after discharge, median (IQR) | 239 (21-809) | 10,245 | 0 |
| Blood transfusion | 3,080 (29.8) | 10,342 | 0 |
| Number of blood transfusions, median (IQR) | 4 (2-8) | 3,080 | 0 |
| Blood transfusion received $\geq 4$ units | 2,291 (22.2) | 10,342 | 0 |
| Length of stay, median (IQR, range) | 7 (5-11) | 10,342 | 0 |
| Prolonged length of stay $\geq 8$ days | 4,744 (45.9) | 10,342 | 0 |

Abbreviations. IQR, interquartile range; PE, pulmonary embolism; DVT, deep vein thrombosis

**Table 5.** Association between clinical outcomes and final diagnosis.

|  | Hemodynamic instability* |  |  | 30 day-rebleeding |  |  | Thromboembolism |  |  | In-hospital mortality |  |  |
| --- | --- | --- | --- | --- | --- | --- | --- | --- | --- | --- | --- | --- |
|  | Absent | Present | P | Absent | Present | P | Absent | Present | P | Absent | Present | P |
| Colonic diverticular bleeding | 1,032 (27.4) | 2,014 (30.6) | 0.001 | 325 (8.6) | 1,501 (22.8) | <0.001 | 27 (0.7) | 38 (0.6) | 0.39 | 81 (2.2) | 16 (0.2) | <0.001 |
| Ischemic colitis | 2,860 (30.4) | 186 (19.8) | <0.001 | 1,796 (19.1) | 30 (3.2) | <0.001 | 61 (0.7) | 4 (0.4) | 0.52 | 94 (0.3) | 3 (0.9) | 0.033 |
| Malignancy | 2993 (29.5) | 53 (27.5) | 0.54 | 1,812 (17.9) | 14 (7.2) | <0.001 | 61 (0.6) | 4 (2.1) | 0.033 | 82 (7.8) | 15 (0.9) | <0.001 |
| Infectious colitis | 3,002 (29.4) | 44 (32.80) | 0.387 | 1,823 (17.9) | 3 (2.2) | <0.001 | 64 (0.6) | 1 (0.8) | 0.573 | 94 (2.2) | 3 (0.9) | 0.131 |
| IBD | 2,977 (29.4) | 69 (32.9) | 0.274 | 1,809 (17.9) | 17 (8.1) | <0.001 | 64 (0.6) | 1 (0.5) | 1 | 97 (1.0) | 0 | 0.27 |
| Radiation colitis | 3,035 (29.5) | 11 (16.7) | 0.022 | 1,814 (17.7) | 12 (18.2) | 0.911 | 65 (0.6) | 0 | 1 | 97 (0.9) | 0 | 1 |
| Other colitis | 3,010 (29.4) | 36 (30.8) | 0.753 | 1,816 (17.8) | 10 (98.6) | 0.009 | 65 (0.6) | 0 | 1 | 95 (0.9) | 2 (1.7) | 0.3 |
| Colorectal angioectasia | 3,010 (29.5) | 36 (27.1) | 0.544 | 1,803 (17.7) | 23 (17.3) | 0.912 | 64 (0.6) | 1 (0.8) | 0.57 | 95 (0.9) | 2 (1.5) | 0.355 |
| Rectal ulcer | 2,966 (29.4) | 80 (31.1) | 0.551 | 1,773 (17.6) | 48 (18.7) | 0.664 | 63 (0.6) | 2 (0.8) | 0.677 | 84 (0.8) | 13 (5.1) | <0.001 |
| Hemorrhoids | 2,991 (29.4) | 55 (29.9) | 0.895 | 1,813 (17.9) | 13 (7.1) | <0.001 | 63 (0.6) | 2 (1.1) | 0.322 | 97 (1.0) | 0 | 0.422 |
| Post-endoscopy bleeding | 2,934 (29.7) | 112 (24.2) | 0.011 | 1,802 (18.2) | 24 (5.2) | <0.001 | 65 (0.7) | 0 | 0.12 | 97 (1.0) | 0 | 0.023 |
| Small bowel bleeding | 2,962 (29.3) | 84 (34.2) | 0.102 | 1,771 (17.5) | 55 (22.4) | 0.05 | 62 (0.6) | 3 (1.2) | 0.201 | 93 (0.9) | 4 (1.6) | 0.295 |
| UGIB | 2,959 (29.0) | 87 (56.9) | <0.001 | 1,821 (17.9) | 5 (3.3) | <0.001 | 61 (0.6) | 4 (2.6) | 0.016 | 89 (0.9) | 8 (5.2) | <0.001 |
| Others | 2,867 (29.6) | 179 (26.7) | 0.108 | 1,755 (18.2) | 71 (10.6) | <0.001 | 60 (0.6) | 5 (0.8) | 0.612 | 66 (0.7) | 31 (4.6) | <0.001 |

|  | Endoscopic therapy need |  |  | Surgery need |  |  | IVR need |  |  | Transfusion need |  |  |
| --- | --- | --- | --- | --- | --- | --- | --- | --- | --- | --- | --- | --- |
|  | Absent | Present | P | Absent | Present | P | Absent | Present | P | Absent | Present | P |
| Colonic diverticular bleeding | 935 (24.8) | 2,444 (37.2) | <0.001 | 97 (2.6) | 45 (0.7) | <0.001 | 41 (1.1) | 176 (2.7) | <0.001 | 974 (25.9) | 2,106 (32.0) | <0.001 |
| Ischemic colitis | 3,374 (35.9) | 5 (0.5) | <0.001 | 135 (1.4) | 7 (0.7) | 0.082 | 216 (2.3) | 1 (0.1) | <0.001 | 3,042 (32.4) | 38 (4.0) | <0.001 |
| Malignancy | 3,358 (33.1) | 21 (10.9) | <0.001 | 110 (1.1) | 32 (16.6) | <0.001 | 215 (2.1) | 2 (1.0) | 0.445 | 3,005 (29.6) | 75 (38.9) | 0.005 |
| Infectious colitis | 3,378 (33.1) | 1 (0.8) | <0.001 | 140 (1.4) | 2 (1.5) | 0.707 | 217 (2.1) | 0 | 0.12 | 3,067 (30.1) | 13 (9.7) | <0.001 |
| IBD | 3,371 (33.3) | 8 (3.8) | <0.001 | 136 (1.3) | 6 (2.9) | 0.062 | 215 (2.1) | 2 (1.0) | 0.331 | 3,048 (30.1) | 32 (15.2) | <0.001 |
| Radiation colitis | 3,327 (32.4) | 52 (78.8) | <0.001 | 142 (1.4) | 0 | 1.000 | 217 (2.1) | 0 | 0.65 | 3,61 (29.8) | 19 (28.8) | 0.859 |
| Other colitis | 3,351 (32.8) | 28 (23.9) | 0.043 | 141 (1.4) | 1 (0.9) | 1 | 217 (2.1) | 0 | 0.182 | 3,041 (29.7) | 39 (33.3) | 0.398 |
| Angioectasia | 3,217 (31.9) | 162 (63.0) | <0.001 | 144 (1.4) | 0 | 0.267 | 215 (2.1) | 2 (1.5) | 1.000 | 3,001 (29.4) | 79 (59.4) | <0.001 |
| Rectal ulcer | 3,291 (32.0) | 163 (62.9) | <0.001 | 137 (1.4) | 5 (2.0) | 0.406 | 215 (2.1) | 2 (0.8) | 0.182 | 2,952 (29.3) | 128 (49.8) | <0.001 |
| Hemorrhoids | 3,369 (33.2) | 10 (5.4) | <0.001 | 134 (1.3) | 8 (4.4) | <0.001 | 217 (2.1) | 0 | 0.035 | 3,036 (29.9) | 44 (23.9) | 0.079 |
| Post-endoscopy bleeding | 2,963 (30.0) | 416 (89.9) | <0.001 | 140 (1.4) | 2 (0.4) | 0.097 | 216 (2.2) | 1 (0.2) | 0.001 | 3,041 (30.8) | 39 (8.4) | <0.001 |
| Small bowel bleeding | 3,322 (33.3) | 57 (23.2) | 0.001 | 119 (1.2) | 23 (9.4) | <0.001 | 198 (2.0) | 19 (7.6) | <0.001 | 2,943 (29.2) | 137 (55.7) | <0.001 |
| UGIB | 3,378 (33.2) | 1 (0.7) | <0.001 | 140 (1.4) | 2 (1.3) | 1.000 | 214 (2.1) | 3 (2.0) | 1.000 | 2,993 (29.4) | 87 (56.9) | <0.001 |
| Others | 3,302 (34.1) | 77 (11.5) | <0.001 | 133 (1.4) | 9 (1.3) | 0.945 | 208 (2.2) | 9 (1.3) | 0.207 | 2,836 (29.3) | 244 (36.4) | <0.001 |

Note. For the analysis of the association between bleeding etiologies and outcomes, etiologies that have > 50 cases were included.
\*Hemodynamic instability was defined as initial systolic blood pressure < 90 mm Hg, initial heart rate $\geq 100$ /min, or the presence of syncope.
Abbreviations. IBD, inflammatory bowel disease; UGIB, upper gastrointestinal bleeding.

### Endoscopic therapy and other procedures

To identify the source of bleeding, multiple procedures were often used during peri-endoscopic procedures: bowel prep with PEG for 66.4% of patients, enema for 19.1%, endoscopic cap for 72.1%, and waterjet scope for 77.0% **(Table 3)**. Endoscopic therapy was performed in 30.7% of patients undergoing endoscopy, mostly clipping (63.8%), followed by band ligation (24.2%), coagulation (8.2%), snare ligation (3.9%), and hypertonic saline-epinephrine (HSE) (1.8%). The success rate of endoscopic therapy was 95.7%, and failure of hemostasis was 4.4%. When initial endoscopic treatment failed, clipping (63.6%) was the most commonly used follow-up technique. Identified post-endoscopy complications were 0.1% perforation and 0.03% diverticulitis. IVR and surgery were performed for 1.4%, and 1.0%, respectively. Secondary endoscopic therapy was performed in 39.5% of patients undergoing repeat colonoscopy (**Supplementary Table 4**). The treatment rate for rebleeding was conservative therapy for 53.1%, endoscopic therapy for 40.6%, IVR for 3.6%, and surgery for 1.7% (**Supplementary Table 4**). The treatment rate for second rebleeding was conservative therapy for 44.8%, endoscopic therapy for 42.1%, IVR for 10.0%, and surgery for 3.7% (**Supplementary Table 5**). The procedure rates of IVR and surgery in second rebleeding were significantly (p<0.01) higher than for first rebleeding. Overall procedure rates during hospitalization were endoscopic therapy for 32.7%, IVR for 2.1%, and surgery for 1.4% (**Table 3**).

### Clinical outcomes

In-hospital rebleeding was identified in 15.2% of patients: 10.8% with one episode of rebleeding, 4.4% with two episodes (**Table 4**). After discharge, rebleeding developed in 25.6%: 22.9% had 1-4 episodes of rebleeding; 1.7% had 5-9 episodes; and 1.0% had ≥ 10 episodes. Thromboembolic events occurred in 0.6% of patients, including acute coronary syndrome in 0.2%, cerebrovascular disease in 0.3% and pulmonary embolism/deep vein thrombosis in 0.2%. In-hospital mortality was 0.9%, mainly due to worsening comorbidities and non-bleeding-related deaths. Only 13% of deaths were directly related to GI bleeding. Out-of-hospital mortality was 6.8% with a median follow-up of 239 days after discharge. Blood transfusion was administered in 29.8% of patients, with a median number of 4 units. The median length of stay was 7 days.

### Association between clinical outcomes and bleeding etiologies

The proportions of clinical outcomes differed according to bleeding etiologies (**Table 5**). Etiologies that were more likely to have adverse outcomes included diverticular bleeding, malignancy, rectal ulcer, small bowel bleeding, or UGIB. For example, patients with diverticular bleeding had significantly higher rates of hemodynamic instability, 30-day-rebleeding, need of endoscopic therapy, IVR, and transfusion, but lower rates of death and surgical need than other etiologies. Patients with small bowel bleeding had significantly higher rates of surgery, IVR, and transfusion than other etigologies. Malignancy or UGIB cases had significantly higher rates of thromboembolism and death than those without them, respectively. At presentation, hemodynamic instability was common in 59.7% of patients with UGIB. In contrast, diseases that have relatively favorable outcomes and may not require intensive care after endoscopy were ischemic colitis, infectious colitis, and post-endoscopy bleeding. For instance, patients with ischemic colitis showed significantly lower rates of hemodynamic status, 30-day-rebleeding, death, and need of endoscopic therapy, IVR, and transfusion than those other etiologies.

### Comparison of clinical data between Japan (JP) and the UK study

Differences of clinical data between JP and the UK are shown in **Supplementary Table 7**. In JP, no missing values in data collection regarding diagnosis, procedures, or outcomes were noted (**Supplementary Table 2**), whereas many of the aforementioned items contained missing values in the UK study, such as data for endoscopic therapy in 5.6% (141/2,528), rebleeding in 5.0% (126/2,528), and length of stay in 6.2% (156/2258). Of baseline characteristics, comorbidity index rates of 1 and ≥ 2 scores and median age were similar in both data sets. However, compared to UK data, JP data had higher proportions of age ≥ 60 y, male, presenting tarry stools, hemoglobin value ≤ 7.0 (g/dl), past history of LGIB, NSAID use, and lower proportions of abdominal pain, low-dose aspirin, and warfarin. In diagnosis, compared to the UK, JP had proportions three times higher or more of performing CT (69.0% vs 20.1%), initial colonoscopy (87.8% vs 29.3%), and repeat colonoscopy (19.3% vs 1.7%). The most common source of bleeding was diverticular bleeding in both groups, followed in the UK by hemorrhoids in 12% and malignancies in 6%, and in JP by ischemic colitis in 9% and procedure-related bleeding in 4.4%. Notably, the unknown bleeding etiology rate was four times lower in JP than the UK (5.2% vs 22.8%). Among procedures, the endoscopic therapy rate was more than 10 times higher in JP than the UK (32.7% vs 2.3%), whereas the rates of IVR and surgical need were not significantly different between the two groups. In outcomes, no significant differences were noted in the rate of in-hospital rebleeding, but overall transfusion rate and transfusion received ≥ 4 units were significantly higher in JP than the UK. Occurrence of acute coronary syndrome and cerebrovascular disease were not significant in either group. In-hospital mortality was more than three times higher in the UK than JP (3.4% vs 0.9%). Median length of stay was longer in JP than in the UK.

## Discussion

In this study, we focused on data from patients with acute hematochezia, rather than colorectal/anal bleeding (LGIB), because the details of bleeding sources remain unclear without endoscopy. We constructed an unprecedentedly large-scale data set and provided information of baseline characteristics, etiologies, interventions, and clinical outcomes. In particular, the high endoscopic rates in JP have revealed a much more detailed and accurate picture of bleeding sources, resulting in performance of endoscopic therapy in 1/3 of all cases, much higher than in previous studies[6,18]. Moreover, bleeding etiologies allowed identification of patients at risk for adverse outcomes. These two findings suggest the importance of accurate diagnosis by colonoscopy, as described in guidelines[1,3]. Finally, comparison of large-scale data in the UK and JP revealed a higher rate of in-hospital mortality in the UK[18], but a higher rate of transfusion in JP, despite similar rates of in-hospital rebleeding and thromboembolism.

Compared to the UK data[18], JP had a three times higher proportion of CT, initial colonoscopy, and repeat colonoscopy, enabling accurate identification of etiologies of acute hematochezia. The diagnostic yield of final diagnosis based on initial and secondary endoscopies reached 94.9%, contributing to identification of 48 diseases, as bleeding etiologies. Only 5% of etiologies remained unknown, a lower rate than in the US at 9%[6], the UK at 23%[18], and Spain at 18%[20]. This is partly because endoscopy is readily accessible in JP hospitals[3]. The most common etiology was diverticular bleeding, at 64%, which is consistent with earlier ALGIB reports[18,20], regardless of different proportions of 30% (75/252) in the US[6], 26% (668/2,528) in the UK[18], and 39% (163/415) in Spain[20]. The top 3 diagnoses other than diverticulosis were ischemic colitis, post-endoscopy bleeding, and rectal ulcer in JP, similar to hemorrhoids, ischemic colitis, and post-endoscopic bleeding in the US[6], but not to hemorrhoids, undetermined colitis, and malignancies in the UK[18]. The differences in order and proportions of etiologies between countries are probably due to the difference in the rates of colonoscopy received and in hospital departments managed (e.g., surgery department in the UK vs gastroenterology in JP)[18].

Among bleeding locations, the ascending colon was most commonly identified in SRH on colonoscopy and extravasation on CT, more than twice as often as the sigmoid colon, suggesting the importance of total colonoscopy. If only sigmoidoscopy were performed, 89% of the overall SRH, and endoscopically treatable lesions would have been missed. Compared to prior studies showing 8% to 26% SRH[15,21,22], we found a high rate of 31%, due to the high use of bowel preparation in 84% before endoscopy and of devices to be attached to the endoscope (e.g., cap, 73% and waterjet, 77%) during endoscopy (**Table 3)**. The high SRH identification rate enabled endoscopic treatment in 33%, which is more than 10 times higher than in the UK[18].

Among endoscopic therapies, clipping is a common worldwide treatment, also the most common in JP (64%). In contrast, HSE and coagulation, commonly used in the US[23], were used in 10% or less of cases in JP. Treatment in JP is characterized by the use of novel therapies such as band ligation and snare ligation, which have been scarcely reported in the US or UK[23,24]. In recent years, the evidence that band ligation is more effective in preventing recurrence than clipping has been accumulating[23,24], so it may become the mainstream treatment for ALGIB. The rate of IVR or surgery in JP was less than 5%, which was similar to the UK[18]. Both treatments were performed more frequently at second rebleeding than first rebleeding, suggesting that physicians are prone to performing endoscopy first, even if extravasation on CT is positive, and to choosing IVR/surgery when uncontrolled bleeding is present, which is in compliance with JP and US guidelines[1,3], but not UK guidelines[2].

In-hospital mortality was more than three times higher in the UK than in JP regardless of a similar rate of ≥ 2 comorbidity score[18], and higher rates of being 60 y or older or having hemoglobin < 7 in JP. This was probably because the UK database listed malignancy more than 3 times as often as JP. Another reason was that the rate of endoscopy and endoscopic treatment was much higher in JP than in the UK, which enhances control of continuous or massive or recurrent bleeding. Uncontrolled LGIB episodes have been suggested to increase the risk of death from worsening comorbidities, although bleeding is not the direct cause of death[8]. The reason the length of stay was longer in JP than the UK is that JP tends to monitor the progress of ALGIB patients carefully since they are administered fasting intravenous drip first, with the diet gradually changing to a normal diet[15,25]. In fact, prior LGIB reports in JP show a median hospital stay of 7 days[15,25].

Guidelines state that the value of the colonoscopy for ALGIB is to identify the bleeding etiology and perform hemostasis if indicated[1,3]. However, in addition to this, we hypothesized that colonoscopy could uncover important factors associated with clinical outcomes. Regrettably, prior ALGIB studies have never evaluated the association between endoscopic diagnoses and outcomes[4–7,26]. Significantly, we found that high-risk etiologies of adverse outcomes were diverticular bleeding, malignancy, rectal ulcer, small bowel bleeding, or UGIB among patients with hematochezia. In contrast, low-risk etiologies were ischemic colitis, infectious colitis, or post-endoscopy bleeding. This indicates that accurate identification of bleeding etiology can stratify patients at risk for adverse outcomes, helping physicians determine whether they should give intensive care, or otherwise recommend prompt discharge after endoscopy. In addition, we associated endoscopic diagnosis with initial characteristics: UGIB is associated with hemodynamic instability, and may be indicative of an UGIB source and warrants an upper endoscopy. Although there was no high quality evidence to support this finding, we confirmed the validity of the UK and US guidelines[1,2].

We recognize limitations to this study. First, this was a retrospective study, leading to some missing values in baseline characteristics (e.g., alcohol intake and INR values), potentially a source of bias. However, there were no missing values for diagnosis, procedures, and outcomes in the JP data, and items with missing values and their rates were lower than in the UK prospective study. Second, all participating institutions have relatively large numbers of beds and 24/7 colonoscopy accessible, and it remains unclear whether our results are applicable to non-emergency hospitals with different settings.

In conclusion, we provided useful clinical data for ALGIB management from 10,342 patients with acute hematochezia. Although some data were similar to the UK nationwide data, the Japanese data are characterized by higher rates of endoscopy, CT examination, and lower mortality. A high endoscopic rate detects accurate bleeding etiologies, enabling us to stratify patients at high or low risk of adverse outcomes, emphasizing the high value of colonoscopy.

## Data Availability

All data relevant to the study are included in the article or uploaded as supplementary information.

## Collaborators

CODE BLUE J-study group: full names and affiliations listed in the online **Supplementary Table 1**.

## Contributors

NN was principal investigator of this study. NN designed and conducted the study, interpreted the data, and mainly wrote the paper. KK (Bokuto hospital), AY, AY. JO, TI, TA, NT, YS, TK, NI, TS, MM, AT, KM, KK (Fukuoka University Chikushi Hospital), SF, TU, MF, HS, SS, TN, JH, TF, YK, AM, SK, TM, RG, HF, YF, NG, YT, KN, NM,KN, TK, YS, SF, KK, TM, YK, KM, KW, and MK designed the study, made decisions and definitions of survey items, and interpreted the data. NN and KM performed the statistical analysis. MF, TI, NU, TK, MK provided corrections and advice on the preparation of the paper.

## Acknowledgements

The authors thank Kazuyo Jo, Shiho Kamimura, Sanae Habu, Akiko Takamatsu, Minako Kajihara, and Kenko Yoshida for their help with the data collection and analysis.

## Abbreviations

ALGIB: acute lower gastrointestinal bleeding
CCI: Charlson comorbidity index
CRP: C-reactive protein
CT: computer tomography
DOACs: direct oral anticoagulants
GIB: gastrointestinal bleeding
HSE: hypertonic saline epinephrine
INR: international normalized ratio
IQR: inter quartile range
IVR: interventional radiology
JP: Japan
LGIB: lower gastrointestinal bleeding
LOS: length of hospital stay
NSAIDs: non-steroidal anti-inflammatory drugs
RCTs: randomized controlled trials
SRH: stigmata of recent hemorrhage
UGIB: upper gastrointestinal bleeding
UK: united kingdom
US: united states

