## Supplemental Tables for "Nationwide large-scale data of acute lower gastrointestinal bleeding in Japan uncover detailed etiologies and relevant outcomes: CODE BLUE J-Study": Supplementary Tables.docx

**Title**

**Running Head:** bleeding etiologies and outcomes

***Correspondence:** Naoyoshi Nagata, MD, PhD.

Department of Gastroenterological Endoscopy, Tokyo Medical University, 6-7-1 Nishishinjuku, Shinjuku-ku, Tokyo 160-0023, Japan

**Supplementary Table 1 Affiliations, ethics committee approval number, and number of cases in 49 participating hospitals**

| No | Prefecture | Institutions | Department | Approval number in ethics committee | Period of data collection | Number of cases |
| --- | --- | --- | --- | --- | --- | --- |
| 1 | Tokyo | Tokyo Medical University | Department of Gastroenterological Endoscopy | T20190244 | 2013.01.02-2019.11.13 | 134 |
| 2 | Tokyo | National Center for Global Health and Medicine | Department of Gastroenterology and Hepatology | 3539 | 2010.01.19-2019.06.18 | 1375 |
| 3 | Tokyo | Tokyo Shinagawa Hospital | Department of Gastroenterology | 20-A-04 | 2014.07.28-2019.12.30 | 250 |
| 4 | Tokyo | Nippon Medical School, Graduate School of Medicine | Department of Gastroenterology | B-2020-147 | 2014.10.31-2019.12.23 | 538 |
| 5 | Chiba | Chiba Hokusoh Hospital, Nippon Medical School | Department of Gastroenterology | 802 | 2011.08.28-2019.12.23 | 188 |
| 6 | Saga | Saga Medical Center Koseikan | Department of Gastroenterology | 20-01-01-03 | 2013.01.02-2019.12.30 | 394 |
| 7 | Tokyo | St. Luke’s International University | Department of Gastroenterology | 20-R012 | 2014.01.03-2019.12.29 | 536 |
| 8 | Okayama | Kawasaki Medical School | Division of Endoscopy and Ultrasonography, and Department of Clinical Pathology and Laboratory Medicine | 3890 | 2014.03.27-2018.2.18 | 156 |
| 9 | Okayama | Kawasaki Medical School General Medical Center | Division of Endoscopy and Ultrasonography, and Department of Clinical Pathology and Laboratory Medicine | 3890 | 2012.02.07-2017.12.27 | 88 |
| 10 | Ibaraki | University of Tsukuba | Department of Gastroenterology, and Division of Endoscopic Center | R02-030 | 2013.01.02-2019.12.30 | 134 |
| 11 | Tokyo | Tokyo Metropolitan Bokutoh Hospital | Department of Gastroenterology | 02-024 | 2013.04.02-2019.12.31 | 808 |
| 12 | Kanagawa | Saiseikai Yokohamashi Tobu Hospital | Emergency and Critical Care Center | 20200030 | 2011.01.22-2019.12.17 | 131 |
| 13 | Tokyo | The University of Tokyo | Department of Gastroenterology | 2020067NI | 2010.01.04-2018.08.10 | 542 |
| 14 | Tokyo | Toranomon Hospital | Department of Gastroenterology | 2021 | 2011.04.02-2019.12.04 | 134 |
| 15 | Aichi | Nagoya University Hospital | Department of Endoscopy | 2020-0152 | 2016.01.02-2019.12.27 | 206 |
| 16 | Hiroshima | Hiroshima City Asa Citizens Hospital | Department of Gastroenterology | 02-1-24 | 2010.01.02-2019.12.27 | 517 |
| 17 | Fukuoka | National Hospital Organization Fukuokahigashi Medical Center | Department of Gastroenterology and Hepatology | 2020-臨-2 | 2017.04.02-20.9.12.3 | 107 |
| 18 | Nara | Nara City Hospital | Department of Gastroenterology and Hepatology, and Center for Digestive and Liver Diseases | NCH倫20-8 | 2013.05.02-2019.12.25 | 264 |
| 19 | Niigata | Graduate School of Medical and Dental Sciences, Niigata University | Division of Gastroenterology | 2020-0052 | 2010.8.28-2019.12.18 | 142 |
| 20 | Kanagawa | St Marianna University School of Medicine | Division of Gastroenterology and Hepatology, Department of Internal Medicine | 4802 | 2013.08.12-2019.12.13 | 343 |
| 21 | Oita | Oita University | Department of Gastroenterology | 1845 | 2010.03.06-2019.12.03 | 199 |
| 22 | Tokyo | Tokyo Saiseikai Central Hospital | Department of Internal Medicine | 2020-015-01 | 2017.01.09-2019.12.29 | 120 |
| 23 | Fukuoka | Fukuoka University Hospital | Department of Gastroenterological Endoscopy | U20-05-016 | 2016.03.30-2019.12.12 | 80 |
| 24 | Fukuoka | Fukuoka University Chikushi Hospital | Department of Gastroenterology | C20-052 | 2010.01.05-2017.03.04 | 199 |
| 25 | Osaka | Kitano Hospital, Tazuke Kofukai Medical Research Institute | Department of Gastroenterology and Hepatology | P200500400 | 2010.1.24-2019.12.21 | 586 |
| 26 | Fukuoka | Graduate School of Medical Sciences, Kyushu University | Department of Medicine and Clinical Science | 2020-289 | 2010.05.30-2019.12.25 | 101 |
| 27 | Miyazaki | University of Miyazaki Hospital | Department of Gastroenterology and Hepatology, and Center for Digestive Disease and Division of Endoscopy | 0-0734 | 2010.07.16-2019.10.20 | 142 |
| 28 | Okinawa | University of the Ryukyus Hospital | Department of Endoscopy | 1656 | 2016.01.04-2019.12.08 | 86 |
| 29 | Okinawa | Naha City Hospital | Department of Gastroenterology | 2004a4 | 2018.11.02-2019.12.26 | 121 |
| 30 | Kagoshima | Kagoshima University Graduate School of Medical and Dental Sciences | Digestive and Lifestyle Diseases | 200041疫 | 2019.02.26-2019.10.07 | 7 |
| 31 | Kagoshima | Kagoshima City Hospital | Department of Gastroenterology | 2020-25 | 2019.01.27-2019.12.27 | 45 |
| 32 | Kagoshima | Kagoshima Kouseiren Hospital | Department of Gastroenterology | 215 | 2019.01.03-2019.12.25 | 28 |
| 33 | Kagoshima | Kagoshima Medical Center | Department of Gastroenterology | 2020-22 | 2019.01.01-2019.12.25 | 40 |
| 34 | Kagoshima | Izumi General Medical Center | Department of Gastroenterology | 60 | 2019.01.08-2019.12.26 | 34 |
| 35 | Kagoshima | Kirishima City Medical Association Medical Center | Department of Gastroenterology | 202005 | 2019.01.16-2019.10.12 | 19 |
| 36 | Kagoshima | Kagoshima Prefectural Oshima Hospital | Department of Gastroenterology | 97 | 2018.12.25-2019.12.27 | 23 |
| 37 | Kyoto | National Hospital Organization Kyoto Medical Center | Department of Gastroenterology | 20-020 | 2011.05.31-2019.12.20 | 205 |
| 38 | Fukushima | Fukushima Medical University | Department of Gastroenterology | 一般2020-112 | 2013.12.07-2019.12.26 | 100 |
| 39 | Tokyo | Tokyo Metropolitan Cancer and Infectious Diseases Center Komagome Hospital | Department of Gastroenterology | 2503 | 2010.06.15-2019.12.16 | 205 |
| 40 | Kanagawa | Kitasato University, School of Medicine | Department of Gastroenterology | C20-174 | 2010.10.07-2019.12.26 | 76 |
| 41 | Osaka | Suita Municipal Hospital | Department of Gastroenterology and Hepatology | 2020-研2 | 2011.03.04-2019.12.30 | 88 |
| 42 | Akita | Akita University Graduate School of Medicine | Department of Gastroenterology and Neurology | 2491 | 2010.09.22-2019.05.11 | 69 |
| 43 | Shizuoka | Japanese Red Cross Shizuoka Hospital | Department of Gastroenterology | 2020-06 | 2010.01.08-2019.12.30 | 173 |
| 44 | Aomori | Hirosaki University Hospital | Division of Endoscopy | 2020-32 | 2010.04.16-2019.12.28 | 112 |
| 45 | Kumamoto | Graduate School of Medical Sciences, Kumamoto University | Department of Gastroenterology and Hepatology | 2040 | 2010.01.03-2019.12.22 | 111 |
| 46 | Fukuoka | National Hospital Organization Kyushu Medical Center | Department of Gastroenterology | 20C065 | 2010.4.12-2019.12.23 | 82 |
| 47 | Iwate | Iwate Medical University | Department of Internal Medicine | MH2020-050 | 2015.01.07-2019.7.19 | 98 |
| 48 | Yamaguchi | Shuto General Hospital | Department of Gastroenterology | H31-24 | 2014.07.25-2019.08.19 | 114 |
| 49 | Saitama | National Defense Medical College | Department of Internal Medicine | 4217 | 2010.04.12-2019.12.07 | 92 |

**Supplementary Table 2 Survey items of baseline characteristics, diagnoses, procedures, and clinical outcomes**

| Category | Clinical factors | Definition | Example | Characteristics　 of value | Number of items | Missing value |
| --- | --- | --- | --- | --- | --- | --- |
| Baseline characteristics | Date of data collection |  | 2030/11/11 | Date | 1 | 0 |
| Baseline characteristics | Participating hospitals |  | X hospital | Free comment | 2 | 0 |
| Baseline characteristics | Research number |  | 1 | Free comment | 3 | 0 |
| Baseline characteristics | Case number |  | 1234567 | Free comment | 4 | 0 |
| Baseline characteristics | Date of admission |  | 2019/3/18 | Date | 5 | 0 |
| Baseline characteristics | Number of ambulances | Number of ambulances accepted by each facility per year in 2018. | 3,000 | Continuous value | 6 | 0 |
| Baseline characteristics | 24/7 colonoscopy | Facilities with a physician who can perform colonoscopy 24 hours a day | 1 | Categorical variable | 7 | 0 |
| Baseline characteristics | Date of onset of hematochezia | Enter the same date if it is the same as the information on admission | 2019/7/5 | Date | 8 | 0 |
| Baseline characteristics | Date of discharge |  | 2019/7/5 | Date | 9 | 0 |
| Baseline characteristics | Date of fasting |  | 2019/12/16 | Date | 10 | 0 |
| Baseline characteristics | Date of starting diet |  | 2019/12/12 | Date | 11 | 0 |
| Baseline characteristics | Age | number, unknown, NA | 86 | Continuous value | 12 | 0 |
| Baseline characteristics | Sex | female, 0; male, 1 | 1 | Categorical variable | 13 | 0 |
| Baseline characteristics | Blood type | Type　O,　O;　 type　A, A; type　B, B; type　AB, 　AB | A | Categorical variable | 14 | 510 (4.8) |
| Baseline characteristics | Height | rounding off, unknown, NA | 162 | Continuous value | 15 | 572 (5.4) |
| Baseline characteristics | Body weight | first decimal place, unknown, NA | 53.3 | Continuous value | 16 | 438 (4.2) |
| Baseline characteristics | Alcohol | current drinker, 1; non-drinker or ever drinker, 0; unknown, NA | 0 | Categorical variable | 17 | 1,465 (13.9) |
| Baseline characteristics | Smoking | never, 0; current smoking, 1; ever, 2 | 2 | Categorical variable | 18 | 1,197 (11.3) |
| Baseline characteristics | Performance status | asymptomatic (Fully active, able to carry on all predisease activities without restriction), 0; Symptomatic but completely ambulatory (Restricted in physically strenuous activity but ambulatory and able to carry out work of a light or sedentary nature. For example, light housework, office work), 1; symptomatic, <50% in bed during the day (Ambulatory and capable of all selfcare but unable to carry out any work activities. Up and about more than 50% of waking hours), 2; symptomatic, >50% in bed, but not bedbound (Capable of only limited self-care, confined to bed or chair 50% or more of waking hours), 3; bedbound (Completely disabled. Cannot carry on any self-care. Totally confined to bed or chair), 4; unknown, NA | 4 | Categorical variable | 19 | 126 (1.2) |
| Baseline characteristics | Systolic blood pressure (mmHg) | number, unknown, NA | 189 | Continuous value | 20 | 209 (2.0) |
| Baseline characteristics | Heart rate (/minutes) | number, unknown, NA | 95 | Continuous value | 21 | 230 (2.2) |
| Baseline characteristics | Syncope/ loss of consciousness | presence, 1; absence, 0; unknown, NA | 0 | Categorical variable | 22 | 19 (0.2) |
| Baseline characteristics | Abdominal pain | presence, 1; absence, 0; unknown, NA | 0 | Categorical variable | 23 | 20 (0.2) |
| Baseline characteristics | Fever | presence, 1 (e.g., >37.0℃) ; absence, 0; unknown, NA | 0 | Categorical variable | 24 | 23 (0.2) |
| Baseline characteristics | Diarrhea | presence, 1; absence, 0; unknown, NA | 0 | Categorical variable | 25 | 36 (0.3) |
| Baseline characteristics | Hematochezia | presence, 1; absence, 0; unknown, NA | 1 | Categorical variable | 26 | 0 |
| Baseline characteristics | Tarry stool | presence, 1; absence, 0; unknown, NA | 1 | Categorical variable | 27 | 22 (0.2) |
| Baseline characteristics | Hemoglobin (g/dl) | first exam date after visiting the hospital, unknown, NA | 7.1 | Continuous value | 28 | 14 (0.1) |
| Baseline characteristics | White blood cell (/ul) | first exam date after visiting the hospital, unknown, NA | 5,930 | Continuous value | 29 | 13 (0.1) |
| Baseline characteristics | Platelet count (/ul) | first exam date after visiting the hospital, unknown, NA | 22.2 | Continuous value | 30 | 17 (0.2) |
| Baseline characteristics | Albumin (/dl) | first exam date after visiting the hospital, unknown, NA | 2.6 | Continuous value | 31 | 503 (4.8) |
| Baseline characteristics | PT-INR | first exam date after visiting the hospital, unknown, NA | 1.22 | Continuous value | 32 | 1,367 (13.0) |
| Baseline characteristics | Hematocrit (%) | first exam date after visiting the hospital, unknown, NA | 21.7 | Continuous value | 33 | 29 (0.3) |
| Baseline characteristics | Blood urea nitrogen (mg/dl) | first exam date after visiting the hospital, unknown, NA | 62.0 | Continuous value | 34 | 80 (0.8) |
| Baseline characteristics | Creatinine (mg/dl) | first exam date after visiting the hospital, unknown, NA | 1.09 | Continuous value | 35 | 84 (0.8) |
| Baseline characteristics | CRP (mg/dl) | first exam date after visiting the hospital, unknown, NA | 5.9 | Continuous value | 36 | 285 (2.7) |
| Baseline characteristics | History of bowel resection | presence, 1; absence, 0; unknown, NA | 0 | Categorical variable | 37 | 4 (0.04) |
| Baseline characteristics | History of chemotherapy | presence, 1; absence, 0; unknown, NA | 0 | Categorical variable | 38 | 26 (0.2) |
| Baseline characteristics | History/ treatment of radiation | presence, 1; absence, 0; unknown, NA | 0 | Categorical variable | 39 | 20 (0.2) |
| Baseline characteristics | History of angiectasia | presence, 1; absence, 0; unknown, NA | 0 | Categorical variable | 40 | 3 (0.03) |
| Baseline characteristics | History of IBD | presence, 1; absence, 0; unknown, NA | 0 | Categorical variable | 41 | 3 (0.03) |
| Baseline characteristics | History of diverticular bleeding | presence, 1; absence, 0; unknown, NA | 0 | Categorical variable | 42 | 10 (0.1) |
| Baseline characteristics | History of ischemic colitis | presence, 1; absence, 0; unknown, NA | 0 | Categorical variable | 43 | 3 (0.03) |
| Baseline characteristics | Diabetes mellitus, uncomplicated | presence, 1; absence, 0; unknown, NA | 0 | Categorical variable | 44 | 2 (0.02) |
| Baseline characteristics | Diabetes mellitus, end-organ damage | presence, 1 (e.g., retinitis, nephropathy and retinopathy); absence, 0; unknown, NA | 0 | Categorical variable | 45 | 2 (0.02) |
| Baseline characteristics | Hemiplegia | presence, 1; absence, 0; unknown, NA | 1 | Categorical variable | 46 | 10 (0.1) |
| Baseline characteristics | CVA or TIA | presence, 1; absence, 0; unknown, NA | 0 | Categorical variable | 47 | 4 (0.04) |
| Baseline characteristics | COPD | presence, 1; absence, 0; unknown, NA | 0 | Categorical variable | 48 | 2 (0.02) |
| Baseline characteristics | Dementia | presence, 1 (e.g., disorder of memory, apraxia, alogia and agnosis, interfere with social life); absence, 0; unknown, NA | 0 | Categorical variable | 49 | 10 (0.1) |
| Baseline characteristics | Connective tissue disease | presence, 1 (e.g., RA, SLE, SJS, MCTD); absence, 0; unknown, NA | 0 | Categorical variable | 50 | 2 (0.02) |
| Baseline characteristics | Myocardial infarction | presence, 1; absence, 0; unknown, NA | 0 | Categorical variable | 51 | 2 (0.02) |
| Baseline characteristics | Chronic heart failure | presence, 1; absence, 0; unknown, NA | 0 | Categorical variable | 52 | 4 (0.04) |
| Baseline characteristics | Peptic ulcer disease | presence, 1 (e.g., gastric ulcer, duodenal ulcer); absence, 0; unknown, NA | 0 | Categorical variable | 53 | 2 (0.02) |
| Baseline characteristics | Moderate CKD | presence, 1 (e.g., eGFR<60, creatinine >3 mg/dL (27 mmol/L)); absence, 0; unknown, NA | 0 | Categorical variable | 54 | 4 (0.04) |
| Baseline characteristics | Severe CKD | presence, 1 (e.g., on dialysis, status post-kidney); absence, 0; unknown, NA | 0 | Categorical variable | 55 | 2 (0.02) |
| Baseline characteristics | Peripheral vascular disease | presence, 1 (e.g., peripheral arterial disease, ASO and TAO); absence, 0; unknown, NA | 0 | Categorical variable | 56 | 2 (0.02) |
| Baseline characteristics | Leukemia/ myeloma | presence, 1; absence, 0; unknown, NA | 0 | Categorical variable | 57 | 2 (0.02) |
| Baseline characteristics | AIDS | presence, 1 (e.g., HIV positive with AIDS index or CD4<100); absence, 0; unknown, NA | 0 | Categorical variable | 58 | 11 (0.1) |
| Baseline characteristics | Solid tumor, localized | presence, 1; absence, 0; unknown, NA | 0 | Categorical variable | 59 | 9 (0.1) |
| Baseline characteristics | Solid tumor, metastatic | presence, 1; absence, 0; unknown, NA | 0 | Categorical variable | 60 | 2 (0.02) |
| Baseline characteristics | Liver disease, mild | presence, 1 (e.g., mild: chronic hepatitis (or cirrhosis without portal hypertension)); absence, 0; unknown, NA | 0 | Categorical variable | 61 | 3 (0.03) |
| Baseline characteristics | Liver disease, moderate to severe | presence, 1 (e.g., Severe: cirrhosis and portal hypertension with variceal bleeding history, moderate = cirrhosis and portal hypertension but no variceal bleeding); absence, 0; unknown, NA | 0 | Categorical variable | 62 | 3 (0.03) |
| Baseline characteristics | Malignant lymphoma | presence, 1; absence, 0; unknown, NA | 0 | Categorical variable | 63 | 2 (0.02) |
| Baseline characteristics | Hypertension | presence, 1 (e.g., drug user); absence, 0; unknown, NA | 0 | Categorical variable | 64 | 2 (0.02) |
| Baseline characteristics | Dyslipidemia | presence, 1 (e.g., drug user); absence, 0; unknown, NA | 0 | Categorical variable | 65 | 3 (0.03) |
| Baseline characteristics | NSAIDs | prior drug use (e.g., Loxoprofen, Diclofenac, Ibuprofen, Zaltoprofen, Mefenamic, Etodolac, Indomethacin) within 1 month | 1 | Categorical variable | 66 | 2 (0.02) |
| Baseline characteristics | COX-2 selective inhibitors | prior drug use within 1 month | 0 | Categorical variable | 67 | 2 (0.02) |
| Baseline characteristics | Low-dose aspirin | prior drug use (e.g., Aspirin) within 1 month | 0 | Categorical variable | 68 | 2 (0.02) |
| Baseline characteristics | Thienopyridine | prior drug use (e.g., Clopidogrel, Prasugrel, Ticlopidine) within 1 month | 0 | Categorical variable | 69 | 2 (0.02) |
| Baseline characteristics | Cilostazol | prior drug use within 1 month | 0 | Categorical variable | 70 | 2 (0.02) |
| Baseline characteristics | Other antiplatelet drug | prior drug use (e.g., Dipyridamole, Beraprost, etc.) within 1 month | 0 | Categorical variable | 71 | 2 (0.02) |
| Baseline characteristics | Warfarin | prior drug use within 1 month | 0 | Categorical variable | 72 | 2 (0.02) |
| Baseline characteristics | DOACs | prior drug use (e.g., Rivaroxaban, Apixaban, Dabigatran, Edoxaban) within 1 month | 0 | Categorical variable | 73 | 2 (0.02) |
| Baseline characteristics | Acetaminophen | prior drug use (e.g., Acetaminophen, Paracetamol, etc.) within 1 month | 0 | Categorical variable | 74 | 2 (0.02) |
| Baseline characteristics | Corticosteroid | prior drug use (e.g., Prednisolone, Dexamethasone, Betamethasone, etc.) within 1 month | 0 | Categorical variable | 75 | 2 (0.02) |
| Diagnosis | CT of the abdomen/pelvis | presence, 1; absence, 0; unknown, NA | 1 | Categorical variable | 76 | 0 |
| Diagnosis | Contrast enhanced CT | presence, 1; absence, 0; unknown, NA | 1 | Categorical variable | 77 | 0 |
| Diagnosis | Urgent CT | over 24h after visiting the hospital, 0; within 24h after visiting the hospital, 1; no exam, NA | 1 | Categorical variable | 78 | 0 |
| Diagnosis | Time to CT scan | time after visiting the hospital | 3 | Continuous value | 79 | 0 |
| Diagnosis | Date of CT examination |  | 2019/3/18 | Date | 80 | 0 |
| Diagnosis | Extravasation on CT | presence, 1; absence, 0; unknown, NA | 1 | Categorical variable | 81 | 0 |
| Diagnosis | Location of extravasation | ileum, 1; cecum, 2; ascending, 3; transverse, 4; descending, 5; sigmoid, 6; rectum, 7 | 3 | Categorical variable | 82 | 0 |
| Diagnosis | Colonic diverticular bleeding on CT | presence, 1; absence, 0; unknown, NA | 1 | Categorical variable | 83 | 0 |
| Diagnosis | Enterocolitis on CT | presence, 1; absence, 0; unknown, NA | 1 | Categorical variable | 84 | 0 |
| Diagnosis | Tumor lesion on CT | presence, 1 (e.g., colorectal cancer, metastasis tumor, etc.); absence, 0; unknown, NA | 1 | Categorical variable | 85 | 0 |
| Diagnosis | Other diagnosis on CT | presence, 1; absence, 0; unknown, NA | 1 | Categorical variable | 86 | 0 |
| Diagnosis | Name of other diagnosis on CT | free comment if other diagnosis is present | Rectal ulcer | Free comment | 87 | 0 |
| Diagnosis | Colonoscopic examination | presence, 1; absence, 0; unknown, NA | 1 | Categorical variable | 88 | 0 |
| Diagnosis | Date of colonoscopy | first date of endoscopy, including the first day of hospitalization | 2019/12/09 | Date | 89 | 0 |
| Diagnosis | Time to colonoscopy | time after visiting the hospital (rounding off) | 3 | Continuous value | 90 | 0 |
| Diagnosis | SRH on endoscopy | presence, 1; absence, 0; unknown, NA | 1 | Categorical variable | 91 | 0 |
| Diagnosis | SRH type | active bleeding, 1; visible vessel, 2; adherent Clot, 3 | 2 | Categorical variable | 92 | 0 |
| Diagnosis | SRH location | ileum, 1; cecum, 2; ascending, 3; transverse, 4; descending, 5; sigmoid, 6; rectum, 7 | 3 | Categorical variable | 93 | 0 |
| Diagnosis | Diverticular bleeding (definitive) | presence, 1; absence, 0; unknown, NA | 1 | Categorical variable | 94 | 0 |
| Diagnosis | Diverticular bleeding (presumptive) | presence, 1; absence, 0; unknown, NA | 0 | Categorical variable | 95 | 0 |
| Diagnosis | Ischemic colitis | presence, 1; absence, 0; unknown, NA | 0 | Categorical variable | 96 | 0 |
| Diagnosis | Post-endoscopic procedure bleeding | absence, 0; after ESD, 1; after polypectomy, 2; after EMR, 3; other hemostatic, 4 | 0 | Categorical variable | 97 | 0 |
| Diagnosis | Rectal ulcer | presence, 1; absence, 0; unknown, NA | 0 | Categorical variable | 98 | 0 |
| Diagnosis | IBD | presence, 1; absence, 0; unknown, NA | 0 | Categorical variable | 99 | 0 |
| Diagnosis | Hemorrhoid bleeding | presence, 1; absence, 0; unknown, NA | 0 | Categorical variable | 100 | 0 |
| Diagnosis | Angiectasia | presence, 1; absence, 0; unknown, NA | 0 | Categorical variable | 101 | 0 |
| Diagnosis | Colorectal cancer | presence, 1; absence, 0; unknown, NA | 0 | Categorical variable | 102 | 0 |
| Diagnosis | Metastatic tumor | presence, 1; absence, 0; unknown, NA | 0 | Categorical variable | 103 | 0 |
| Diagnosis | Other tumor | presence, 1; absence, 0; unknown, NA | 0 | Categorical variable | 104 | 0 |
| Diagnosis | Polyp | presence, 1; absence, 0; unknown, NA | 0 | Categorical variable | 105 | 0 |
| Diagnosis | Infectious colitis | presence, 1; absence, 0; unknown, NA | 0 | Categorical variable | 106 | 0 |
| Diagnosis | Radiation colitis | presence, 1; absence, 0; unknown, NA | 0 | Categorical variable | 107 | 0 |
| Diagnosis | Non-specific colitis | presence, 1; absence, 0; unknown, NA | 0 | Categorical variable | 108 | 0 |
| Diagnosis | Drug-induced ulcer | presence, 1; absence, 0; unknown, NA | 0 | Categorical variable | 109 | 0 |
| Diagnosis | Non-specific ulcer | presence, 1; absence, 0; unknown, NA | 0 | Categorical variable | 110 | 0 |
| Diagnosis | Colorectal varix | presence, 1; absence, 0; unknown, NA | 0 | Categorical variable | 111 | 0 |
| Diagnosis | Dieulafoy ulcer | presence, 1; absence, 0; unknown, NA | 0 | Categorical variable | 112 | 0 |
| Diagnosis | Postoperative anastomotic bleeding | presence, 1; absence, 0; unknown, NA | 0 | Categorical variable | 113 | 0 |
| Diagnosis | Meckel's diverticular bleeding | presence, 1; absence, 0; unknown, NA | 0 | Categorical variable | 114 | 0 |
| Diagnosis | Anal lesion | presence, 1; absence, 0; unknown, NA | 0 | Categorical variable | 115 | 0 |
| Diagnosis | Diverticulitis | presence, 1; absence, 0; unknown, NA | 0 | Categorical variable | 116 | 0 |
| Diagnosis | Small bowel bleeding (definitive) | presence, 1; absence, 0; unknown, NA | 0 | Categorical variable | 117 | 0 |
| Diagnosis | Small bowel bleeding (presumptive) | presence, 1; absence, 0; unknown, NA | 0 | Categorical variable | 118 | 0 |
| Diagnosis | UGIB | presence, 1; absence, 0; unknown, NA | 0 | Categorical variable | 119 | 0 |
| Diagnosis | Name of other diagnosis | free comment | 0 | Free comment | 120 | 0 |
| Diagnosis | Unknown | presence, 1; absence, 0; unknown, NA | 0 | Categorical variable | 121 | 0 |
| Diagnosis | 2nd　colonoscopic examination | presence, 1; absence, 0; unknown, NA | 1 | Categorical variable | 122 | 0 |
| Diagnosis | Date of 2nd colonoscopy | first date of endoscopy, including the first day of hospitalization | 2019/12/09 | Date | 123 | 0 |
| Diagnosis | Time to 2nd colonoscopy | time after visiting the hospital (rounding off) | 3 | Categorical variable | 124 | 0 |
| Diagnosis | SRH on 2nd endoscopy | presence, 1; absence, 0; unknown, NA | 1 | Categorical variable | 125 | 0 |
| Diagnosis | SRH type on 2nd endoscopy | active bleeding, 1; exposed vessel, 2; adherent Clot, 3 | 2 | Categorical variable | 126 | 0 |
| Diagnosis | SRH location on 2nd endoscopy | ileum, 1; cecum, 2; ascending, 3; transverse, 4; descending, 5; sigmoid, 6; rectum, 7 | 3 | Categorical variable | 127 | 0 |
| Diagnosis | Diverticular bleeding (definitive) at 2nd endoscopy | presence, 1; absence, 0; unknown, NA | 1 | Categorical variable | 128 | 0 |
| Diagnosis | Diverticular bleeding (presumptive) at 2nd endoscopy | presence, 1; absence, 0; unknown, NA | 0 | Categorical variable | 129 | 0 |
| Diagnosis | Ischemic colitis at 2nd endoscopy | presence, 1; absence, 0; unknown, NA | 0 | Categorical variable | 130 | 0 |
| Diagnosis | Post-endoscopic procedure bleeding at 2nd endoscopy | absence, 0; after ESD, 1; after polypectomy, 2; after EMR, 3; other hemostatic, 4 | 2 | Categorical variable | 131 | 0 |
| Diagnosis | Rectal ulcer at 2nd endoscopy | presence, 1; absence, 0; unknown, NA | 0 | Categorical variable | 132 | 0 |
| Diagnosis | IBD at 2nd endoscopy | presence, 1; absence, 0; unknown, NA | 0 | Categorical variable | 133 | 0 |
| Diagnosis | Hemorrhoid bleeding at 2nd endoscopy | presence, 1; absence, 0; unknown, NA | 0 | Categorical variable | 134 | 0 |
| Diagnosis | Angiectasia at 2nd endoscopy | presence, 1; absence, 0; unknown, NA | 0 | Categorical variable | 135 | 0 |
| Diagnosis | Colorectal cancer at 2nd endoscopy | presence, 1; absence, 0; unknown, NA | 0 | Categorical variable | 136 | 0 |
| Diagnosis | Metastatic tumor at 2nd endoscopy | presence, 1; absence, 0; unknown, NA | 0 | Categorical variable | 137 | 0 |
| Diagnosis | Other tumor at 2nd endoscopy | presence, 1; absence, 0; unknown, NA | 0 | Categorical variable | 138 | 0 |
| Diagnosis | Polyp at 2nd endoscopy | presence, 1; absence, 0; unknown, NA | 0 | Categorical variable | 139 | 0 |
| Diagnosis | Infectious colitis at 2nd endoscopy | presence, 1; absence, 0; unknown, NA | 0 | Categorical variable | 140 | 0 |
| Diagnosis | Radiation colitis at 2nd endoscopy | presence, 1; absence, 0; unknown, NA | 0 | Categorical variable | 141 | 0 |
| Diagnosis | Non-specific colitis at 2nd endoscopy | presence, 1; absence, 0; unknown, NA | 0 | Categorical variable | 142 | 0 |
| Diagnosis | Drug-induced ulcer at 2nd endoscopy | presence, 1; absence, 0; unknown, NA | 0 | Categorical variable | 143 | 0 |
| Diagnosis | Non-specific ulcer at 2nd endoscopy | presence, 1; absence, 0; unknown, NA | 0 | Categorical variable | 144 | 0 |
| Diagnosis | Colorectal varix at 2nd endoscopy | presence, 1; absence, 0; unknown, NA | 0 | Categorical variable | 145 | 0 |
| Diagnosis | Dieulafoy ulcer at 2nd endoscopy | presence, 1; absence, 0; unknown, NA | 0 | Categorical variable | 146 | 0 |
| Diagnosis | Postoperative anastomotic bleeding at 2nd endoscopy | presence, 1; absence, 0; unknown, NA | 0 | Categorical variable | 147 | 0 |
| Diagnosis | Meckel's diverticular bleeding at 2nd endoscopy | presence, 1; absence, 0; unknown, NA | 0 | Categorical variable | 148 | 0 |
| Diagnosis | Anal lesion at 2nd endoscopy | presence, 1; absence, 0; unknown, NA | 0 | Categorical variable | 149 | 0 |
| Diagnosis | Diverticulitis at 2nd endoscopy | presence, 1; absence, 0; unknown, NA | 0 | Categorical variable | 150 | 0 |
| Diagnosis | Small bowel bleeding (definitive) at 2nd endoscopy | presence, 1; absence, 0; unknown, NA | 0 | Categorical variable | 151 | 0 |
| Diagnosis | Small bowel bleeding (presumptive) at 2nd endoscopy | presence, 1; absence, 0; unknown, NA | 0 | Categorical variable | 152 | 0 |
| Diagnosis | UGIB at 2nd endoscopy | presence, 1; absence, 0; unknown, NA | 0 | Categorical variable | 153 | 0 |
| Diagnosis | Name of other diagnosis at 2nd endoscopy | free comment | 0 | Free comment | 154 | 0 |
| Diagnosis | Unknown at 2nd endoscopy | presence, 1; absence, 0; unknown, NA | 0 | Categorical variable | 155 | 0 |
| Procedures | Type of bowel prep | PEG, 1; PEG + enema, 2; enema, 3 | 1 | Categorical variable | 156 | 0 |
| Procedures | Endoscopic cap use | absence, 0; long, 1; short, 2; ST　hood, 3; other, 4 | 0 | Categorical variable | 157 | 0 |
| Procedures | Waterjet scope use | waterjet scope use or use of waterjet in view of endoscopic image, 1; absence, 0; unknown, NA | 1 | Categorical variable | 158 | 0 |
| Procedures | PEG use in water jet scope | PEG in waterjet presence, 1; absence, 0; unknown, NA | 0 | Categorical variable | 159 | 0 |
| Procedures | Conservative therapy | presence, 1; absence, 0; unknown, NA | 0 | Categorical variable | 160 | 0 |
| Procedures | Endoscopic therapy | presence, 1; absence, 0; unknown, NA | 1 | Categorical variable | 161 | 0 |
| Procedures | Secondary endoscopic therapy | indirect clipping, 1; direct clipping, 2; band ligation, 3; Snare ligation,4; HSE, 5; OTSC, 6; coagulation,7; other therapy, 8 | 1 | Categorical variable | 162 | 0 |
| Procedures | Additional comment of endoscopic therapy | free comment | 0 | Free comment | 163 | 0 |
| Procedures | Success of endoscopic therapy | completion, 1; no completion, 2 | 0 | Categorical variable | 164 | 0 |
| Procedures | Type of treatment of failure | clipping, 1; band, 2; HSE, 3; OTSC, 4; coagulation, 5; other therapy, 6 | 1 | Categorical variable | 165 | 0 |
| Procedures | Post-endoscopy perforation or diverticulitis | no complication, 0; perforation, 1; diverticulitis, 2 (imaging diagnosis with abdominal pain and/or fever); other, 3 | 1 | Categorical variable | 166 | 0 |
| Procedures | Location of perforation/ diverticulitis | ileum, 1; cecum, 2; ascending, 3; transverse, 4; descending, 5; sigmoid, 6; rectum, 7 | 3 | Categorical variable | 167 | 0 |
| Procedures | IVR/ TAE | presence, 1; absence, 0; unknown, NA | 0 | Categorical variable | 168 | 0 |
| Procedures | Surgery | presence, 1; absence, 0; unknown, NA | 0 | Categorical variable | 169 | 0 |
| Procedures | Barium impaction therapy | presence, 1; absence, 0; unknown, NA | 0 | Categorical variable | 170 | 0 |
| Procedures | Type of bowel prep for 2nd endoscopy | PEG, 1; PEG + enema, 2; enema, 3 | 1 | Categorical variable | 171 | 0 |
| Procedures | Endoscopic cap use for 2nd endoscopy | absence, 0; long, 1; short, 2; ST hood, 3; other, 4 | 0 | Categorical variable | 172 | 0 |
| Procedures | Waterjet scope use for 2nd endoscopy | waterjet scope use or use of waterjet in view of endoscopic image, 1; absence, 0; unknown, NA | 1 | Categorical variable | 173 | 0 |
| Procedures | PEG use in water jet scope for 2nd endoscopy | PEG in waterjet presence, 1; absence, 0; unknown, NA | 1 | Categorical variable | 174 | 0 |
| Procedures | 2nd 　conservative therapy | presence, 1; absence, 0; unknown, NA | 0 | Categorical variable | 175 | 0 |
| Procedures | 2nd 　endoscopic therapy | presence, 1; absence, 0; unknown, NA | 1 | Categorical variable | 176 | 0 |
| Procedures | Type of treatment at 2nd endoscopy | indirect clipping, 1; direct clipping, 2; band ligation, 3; Snare ligation,4; HSE, 5; OTSC, 6; coagulation,7; other therapy, 8 | 1 | Categorical variable | 177 | 0 |
| Procedures | Additional comment of therapy at 2nd endoscopy | free comment | 0 | Free comment | 178 | 0 |
| Procedures | Success of therapy at 2nd endoscopy | completion, 1; no completion ,2 | 0 | Categorical variable | 179 | 0 |
| Procedures | Type of treatment of failure at 2nd endoscopy | clipping, 1; band, 2; HSE, 3; OTSC, 4; coagulation, 5; other therapy, 6 | 1 | Categorical variable | 180 | 0 |
| Procedures | Post-2nd endoscopy perforation or diverticulitis | no complication, 0; perforation, 1; diverticulitis, 2 (imaging diagnosis with abdominal pain and/or fever); other, 3 | 1 | Categorical variable | 181 | 0 |
| Procedures | IVR/ TAE after 2nd endoscopy | presence, 1; absence, 0; unknown, NA | 0 | Categorical variable | 182 | 0 |
| Procedures | Surgery after 2nd endoscopy | presence, 1; absence, 0; unknown, NA | 0 | Categorical variable | 183 | 0 |
| Procedures | Barium impaction therapy after 2nd endoscopy | presence, 1; absence, 0; unknown, NA | 0 | Categorical variable | 184 | 0 |
| Procedures | Conservative therapy for rebleeding | presence, 1; absence, 0; unknown, NA | 0 | Categorical variable | 185 | 0 |
| Procedures | Endoscopic therapy for rebleeding | presence, 1; absence, 0; unknown, NA | 1 | Categorical variable | 186 | 0 |
| Procedures | Types of endoscopic therapy for rebleeding | clipping, 1; band ligation, 2; HSE, 3; OTSC, 4; coagulation, 5; other therapy, 6 | 1,3 | Categorical variable | 187 | 0 |
| Procedures | IVR/ TAE for rebleeding | presence, 1; absence, 0; unknown, NA | 0 | Categorical variable | 188 | 0 |
| Procedures | Surgery for rebleeding | presence, 1; absence, 0; unknown, NA | 0 | Categorical variable | 189 | 0 |
| Procedures | Other therapy for rebleeding | free comment | 0 | Free comment | 190 | 0 |
| Procedures | Conservative therapy for 2nd rebleeding | presence, 1; absence, 0; unknown, NA | 0 | Categorical variable | 191 | 0 |
| Procedures | Endoscopic therapy for 2nd rebleeding | presence, 1; absence, 0; unknown, NA | 0 | Categorical variable | 192 | 0 |
| Procedures | Types of endoscopic therapy for 2nd rebleeding | clipping, 1; band ligation, 2; HSE, 3; OTSC, 4; coagulation, 5; other therapy, 6 | 1 | Categorical variable | 193 | 0 |
| Procedures | IVR/ TAE for 2nd rebleeding | presence, 1; absence, 0; unknown, NA | 0 | Categorical variable | 194 | 0 |
| Procedures | Surgery for 2nd rebleeding | presence, 1; absence, 0; unknown, NA | 0 | Categorical variable | 195 | 0 |
| Procedures | Other therapy for 2nd rebleeding | free comment | Barium impaction therapy | Free comment | 196 | 0 |
| Outcomes | Rebleeding | bleeding > 24 h after admission | 1 | Categorical variable | 197 | 0 |
| Outcomes | Date of rebleeding |  | 2019/6/17 | Date | 198 | 0 |
| Outcomes | Overt bleeding at rebleeding | presence, 1 (e.g., hematochezia or bloody stools); absence, 0; unknown, NA | 1 | Categorical variable | 199 | 0 |
| Outcomes | Decreased hemoglobin > 2 (g/dl) at rebleeding | presence, 1 (e.g., decreased by ≥ 2 points compared with the most recent data at the time of rebleeding); absence, 0; unknown, NA | 1 | Categorical variable | 200 | 0 |
| Outcomes | Blood pressure<100 mmHg at rebleeding | presence, 1 (e.g., lowest BP at rebleeding); absence, 0; unknown, NA | 1 | Categorical variable | 201 | 1 (0.01) |
| Outcomes | Heart rate>100 /minute at rebleeding | Presence (highest pulse at rebleeding), 1; absence, 0 | 0 | Categorical variable | 202 | 2 (0.02) |
| Outcomes | 2nd 　rebleeding | bleeding > 24 h after admission | 1 | Categorical variable | 203 | 3 (0.03) |
| Outcomes | 2nd 　date of rebleeding |  | 2019/6/20 | Date | 204 | 0 |
| Outcomes | Overt bleeding at 2nd rebleeding | presence, 1 (e.g., hematochezia or bloody stools); absence, 0; unknown, NA | 1 | Categorical variable | 205 | 0 |
| Outcomes | Decreased hemoglobin > 2 (g/dl) at 2nd rebleeding | presence, 1 (e.g., decreased by ≥ 2 points compared with the most recent data at the time of rebleeding); absence, 0; unknown, NA | 1 | Categorical variable | 206 | 0 |
| Outcomes | Blood pressure<100 mmHg at 2nd rebleeding | presence (e.g., lowest BP at rebleeding), 1; absence, 0; unknown, NA | 1 | Categorical variable | 207 | 0 |
| Outcomes | Heart rate>100 /minute at 2nd rebleeding | (highest pulse at rebleeding), 1; absence, 0 | 1 | Categorical variable | 208 | 0 |
| Outcomes | Occurrence of ACS | presence, 1 (e.g., acute myocardial infarction or angina pectoris); absence, 0; unknown, NA | 0 | Categorical variable | 209 | 0 |
| Outcomes | Occurrence of cerebrovascular disease | presence (e.g. TIA, cerebral infarction), 1; absence, 0; unknown, NA | 0 | Categorical variable | 210 | 0 |
| Outcomes | Occurrence of PE/ DVT | presence, 1; absence, 0; unknown, NA | 0 | Categorical variable | 211 | 0 |
| Outcomes | Occurrence of death | presence, 1; absence, 0; unknown, NA | 1 | Categorical variable | 212 | 0 |
| Outcomes | Direct cause of death | free comment | Pneumoniae | Free comment | 213 | 0 |
| Outcomes | Indirect cause of death | free comment | Pancreatic cancer | Free comment | 214 | 0 |
| Outcomes | Number of blood transfusions | number, unknown, NA | 7 | Continuous value | 215 | 29 (0.3) |
| Outcomes | Date of confirmation of final patient visit after discharge |  | 2019/12/09 | Date | 216 | 0 |
| Outcomes | Date of rebleeding after discharge |  | 2019/12/09 | Date | 217 | 0 |
| Outcomes | Date of death after discharge |  | 2019/12/09 | Date | 218 | 0 |
| Outcomes | Number of rebleedings after discharge | number, unknown, NA | 5 | Continuous value | 219 | 0 |

Abbreviations. IQR, interquartile range; PT-INR, international normalized ratio of prothrombin time; CRP, C-reactive protein; IBD, inflammatory bowel disease; CVA, cerebral vascular accident; TIA, transient ischemic attacks; COPD, chronic obstructive pulmonary disease; RA, rheumatoid arthritis; SLE, systemic lupus erythematosus; SJS, Sjogren's syndrome; MCTD, mixed connective tissue disease; CKD, chronic kidney disease; eGFR, estimated glomerular filtration rate; ASO, arteriosclerosis obliterans; TAO, thromboangiitis obliterans; AIDS, acquired immune deficiency syndrome; HIV, human immunodeficiency virus; CD4, cluster of differentiation 4; NSAIDs, non-steroidal anti-inflammatory drugs; COX, cyclooxygenase; DOAC, direct oral anticoagulants; CT, computer tomography; SRH, Stigmata of recent hemorrhage; ESD, endoscopic submucosal dissection; EMR, endoscopic mucosal resection; IBD, inflammatory bowel disease; UGIB, upper gastrointestinal bleeding; SRH, Stigmata of recent hemorrhage; PEG, polyethylene glycol; HSE, hypertonic saline epinephrine; OTSC, over the scope clip; IVR, interventional radiology; TAE, transarterial arterial embolization; BP, blood pressure; ACS, acute coronary syndrome; PE, pulmonary embolism; DVT, deep vein thrombosis

**Supplementary Table 3 Diagnosis based on first colonoscopy with CT**

| Factor | Value | Number of cases | Missing value |
| --- | --- | --- | --- |
| Diverticular bleeding | 6531 (63.2) | 10,342 | 0 |
| Diverticular bleeding (definitive) | 2020 (19.5) | 10,342 | 0 |
| Diverticular bleeding (presumptive) | 4511 (43.6) | 10,342 | 0 |
| Ischemic colitis | 939 (9.1) | 10,342 | 0 |
| Post-procedure bleeding | 461 (4.5) | 10,342 | 0 |
| Post-procedure bleeding, ESD | 141 (1.4) | 10,342 | 0 |
| Post-procedure bleeding, polypectomy | 73 (0.7) | 10,342 | 0 |
| Post-procedure bleeding, EMR | 220 (2.1) | 10,342 | 0 |
| Post-procedure bleeding, biopsy | 16 (0.2) | 10,342 | 0 |
| Post-procedure bleeding, other therapy | 12 (0.1) | 10,342 | 0 |
| Rectal ulcer | 249 (2.4) | 10,342 | 0 |
| IBD | 208 (2.0) | 10,342 | 0 |
| Hemorrhoid bleeding | 186 (1.8) | 10,342 | 0 |
| Angiectasia | 117 (1.1) | 10,342 | 0 |
| Colorectal cancer | 158 (1.5) | 10,342 | 0 |
| Metastatic tumor | 15 (0.2) | 10,342 | 0 |
| Other tumor | 9 (0.1) | 10,342 | 0 |
| Polyp | 36 (0.4) | 10,342 | 0 |
| Infectious colitis | 134 (1.3) | 10,342 | 0 |
| Radiation colitis | 65 (0.6) | 10,342 | 0 |
| Non-specific colitis | 46 (0.4) | 10,342 | 0 |
| Drug-induced ulcer | 14 (0.1) | 10,342 | 0 |
| Non-specific ulcer | 55 (0.5) | 10,342 | 0 |
| Colorectal varix | 25 (0.2) | 10,342 | 0 |
| Dieulafoy ulcer | 13 (0.1) | 10,342 | 0 |
| Postoperative anastomotic bleeding | 15 (0.2) | 10,342 | 0 |
| Meckel's diverticular bleeding | 9 (0.1) | 10,550 | 0 |
| Anal lesion | 12 (0.1) | 10,342 | 0 |
| Diverticulitis | 7 (0.1) | 10,342 | 0 |
| Small bowel bleeding | 226 (2.2) | 10,342 | 0 |
| Small bowel bleeding (definitive) | 105 (1.0) | 10,342 | 0 |
| Small bowel bleeding (presumptive) | 121 (1.2) | 10,342 | 0 |
| UGIB | 152 (1.5) | 10,342 | 0 |
| Other diagnosis | 39 (0.4) | 10,342 | 0 |
| Unknown | 630 (6.1) | 10,342 | 0 |

Abbreviations. IQR, interquartile range; ESD, endoscopic submucosal dissection; EMR, endoscopic mucosal resection; IBD, inflammatory bowel disease; UGIB, upper gastrointestinal bleeding

**Supplementary Table 4 Endoscopic procedures among patients undergoing second colonoscopy (N=1,992)**

| Factor | Value | Number of cases | Missing value |
| --- | --- | --- | --- |
| Type of bowel prep for 2nd endoscopy, PEG or enema | 1,580 (79.3) | 1,992 | 0 |
| Type of bowel prep for 2nd endoscopy, PEG | 1,314 (66.0) | 1,992 | 0 |
| Type of bowel prep for 2nd endoscopy, enema | 298 (15.0) | 1,992 | 0 |
| Endoscopic cap use for 2nd endoscopy | 1,501 (75.4) | 1,992 | 0 |
| Endoscopic cap use for 2nd endoscopy, long | 389 (19.5) | 1,992 | 0 |
| Endoscopic cap use for 2nd endoscopy, short | 1,105 (55.5) | 1,992 | 0 |
| Endoscopic cap use for 2nd endoscopy, ST hood | 6 (0.3) | 1,992 | 0 |
| Endoscopic cap use for 2nd endoscopy, other | 1 (0.1) | 1,992 | 0 |
| PEG use in water jet scope for 2nd endoscopy | 82 (4.1) | 1,992 | 0 |
| 2nd conservative therapy | 1,151 (57.8) | 1,992 | 0 |
| 2nd endoscopic therapy | 786 (39.5) | 1,992 | 0 |
| Type of treatment at 2nd endoscopy, indirect clipping | 358 (18.0) | 1,992 | 0 |
| Type of treatment at 2nd endoscopy, direct clipping | 133 (6.7) | 1,992 | 0 |
| Type of treatment at 2nd endoscopy, band ligation | 194 (9.7) | 1,992 | 0 |
| Type of treatment at 2nd endoscopy, Snare ligation | 25 (1.3) | 1,992 | 0 |
| Type of treatment at 2nd endoscopy, HSE | 8 (0.4) | 1,992 | 0 |
| Type of treatment at 2nd endoscopy, OTSC | 0 | 1,992 | 0 |
| Type of treatment at 2nd endoscopy, coagulation | 63 (3.2) | 1,992 | 0 |
| Type of treatment at 2nd endoscopy, other endoscopic therapy | 17 (0.9) | 1,992 | 0 |
| Success of therapy at 2nd endoscopy | 725 (36.4) | 1,992 | 0 |
| Type of treatment of failure at 2nd endoscopy, clipping | 37 (1.9) | 1,992 | 0 |
| Type of treatment of failure at 2nd endoscopy, band ligation | 7 (0.4) | 1,992 | 0 |
| Type of treatment of failure at 2nd endoscopy, failure, HSE | 7 (0.4) | 1,992 | 0 |
| Type of treatment of failure at 2nd endoscopy, OTSC | 0 | 1,992 | 0 |
| Type of treatment of failure at 2nd endoscopy, coagulation | 2 (0.1) | 1,992 | 0 |
| Type of treatment of failure at 2nd endoscopy, other therapy | 7 (0.4) | 1,992 | 0 |
| Post-2nd endoscopy perforation | 2 (0.1) | 1,992 | 0 |
| Post-2nd endoscopy diverticulitis | 0 | 1,992 | 0 |
| Post-2nd endoscopy other | 1 (0.1) | 1,992 | 0 |
| IVR after 2nd endoscopy | 19 (0.2) | 10,342 | 0 |
| Surgery after 2nd endoscopy | 17 (0.2) | 10,342 | 0 |
| Barium impaction therapy after 2nd endoscopy | 50 (0.5) | 10,342 | 0 |

Abbreviations. IQR, interquartile range; PEG, polyethylene glycol; HSE, hypertonic saline epinephrine; OTSC, over the scope clip; IVR, interventional radiology.

**Supplementary Table 5 Treatment for rebleeding (N=1,573)**

| Factor | Value | Number of cases | Missing value |
| --- | --- | --- | --- |
| Conservative therapy for rebleeding | 835 (53.1) | 1,573 | 0 |
| Endoscopic therapy for rebleeding | 659 (40.6) | 1,573 | 0 |
| Endoscopic therapy for rebleeding, clipping | 424 (27.0) | 1,573 | 0 |
| Endoscopic therapy for rebleeding, band ligation | 134 (8.5) | 1,573 | 0 |
| Endoscopic therapy for rebleeding, HSE | 28 (1.8) | 1,573 | 0 |
| Endoscopic therapy for rebleeding, OTSC | 2 (0.1) | 1,573 | 0 |
| Endoscopic therapy for rebleeding, coagulation | 34 (2.2) | 1,573 | 0 |
| Endoscopic therapy for rebleeding, other | 37 (2.4) | 1,573 | 0 |
| IVR for rebleeding | 56 (3.6) | 1,573 | 0 |
| Surgery for rebleeding | 27 (1.7) | 1,573 | 0 |
| Other therapy for rebleeding | 41 (2.6) | 1,573 | 0 |

Abbreviations. IQR, interquartile range; HSE, hypertonic saline-epinephrine; OTSC, over the scope clip; IVR, interventional radiology

**Supplementary Table 6 Treatment for second rebleeding (N=458)**

| Factor | Value | Number of cases | Missing value |
| --- | --- | --- | --- |
| Conservative therapy for 2nd rebleeding | 205 (44.8) | 458 | 0 |
| Endoscopic therapy for 2nd rebleeding | 193 (42.1) | 458 | 0 |
| Endoscopic therapy for 2nd rebleeding, clipping | 125 (27.3) | 458 | 0 |
| Endoscopic therapy for 2nd rebleeding, band ligation | 44 (9.6) | 458 | 0 |
| Endoscopic therapy for 2nd rebleeding, HSE | 12 (2.6) | 458 | 0 |
| Endoscopic therapy for 2nd rebleeding, OTSC | 0 | 458 | 0 |
| Endoscopic therapy for 2nd rebleeding, coagulation | 5 (1.1) | 458 | 0 |
| Endoscopic therapy for 2nd rebleeding, other | 12 (2.6) | 458 | 0 |
| IVR for 2nd rebleeding | 46 (10.0) | 458 | 0 |
| Surgery for 2nd rebleeding | 17 (3.7) | 458 | 0 |
| Other therapy for 2nd rebleeding | 12 (2.6) | 458 | 0 |

Abbreviations. IQR, interquartile range; HSE, hypertonic saline-epinephrine; OTSC, over the scope clip; IVR, interventional radiology.

**Supplementary Table 7 Comparison of clinical data between Japan and the UK**

| Japan (n=10,342) |  |  | UK (n=2,528) |  |  |
| --- | --- | --- | --- | --- | --- |
| Factor | Value | Missing value | Value | Missing value | P value |
| Baseline characteristics |  |  |  |  |  |
| Outpatient onset | 10,342 (100) | 0 | 2,331 (92.2) | NA | <0.001 |
| Age | 74 (63-82) | 0 | 74 (57–83) | 0 | NA |
| Age ≥ 60 y | 8,327 (80.5) | 0 | 1,829 (72.3) | 0 | <0.001 |
| Male | 6,317 (61.1) | 0 | 1,209 (48.0) | 7 (0.3) | <0.001 |
| Abdominal pain | 1,664 (16.1) | 19 (0.2) | 492 (19.5) | 6 (0.24) | <0.001 |
| Tarry stools | 593 (5.8) | 21 (0.2) | 120 (4.8) | 6 (0.24) | 0.052 |
| Hemoglobin ≤ 7.0 (g/dl) | 798 (7.7) | 0 | 140 (5.6) | 15 (0.59) | <0.001 |
| Past history LGIB | 3,090 (30.0) | 0 | 408 (20.1) | 495 (19.58) | <0.001 |
| Charlson Comorbidity Index, 0 | 4,124 (39.9) | 0 | 1,066(42.3) |  | 0.035 |
| Charlson Comorbidity Index, 1 | 2,431 (23.5) | 0 | 570 (20.6) |  | 0.307 |
| Charlson Comorbidity Index, ≥ 2 | 3,787 (36.6) | 0 | 885 (35.1) | 7 (0.28) | 0.157 |
| Diabetes mellitus, uncomplicated | 1,933 (18.7) | 0 | 377 (14.9) | NA | <0.001 |
| Cerebral vascular accident or TIA | 1,475 (14.3) | 2 (0.02) | 217 (8.6) | NA | <0.001 |
| COPD | 315 (3.1) | 0 | 298 (11.8) | NA | <0.001 |
| Dementia | 565 (5.5) | 8 (0.1) | 150 (5.9) | NA | 0.359 |
| Myocardial infarction | 418 (4.0) | 0 | 285 (11.3) | NA | <0.001 |
| Chronic heart failure | 1,660 (16.1) | 0 | 159 (6.3) | NA | 0.001 |
| NSAIDs | 1,177 (11.4) | 0 | 146 (5.8) | 0 | <0.001 |
| Low-dose aspirin | 2,056 (20.0) | 0 | 584 (23.1) | 0 | <0.001 |
| Thienopyridine | 1,014 (9.8) | 0 | 235 (9.3) | 0 | 0.439 |
| Number of antiplatelet drugs, 2 | 660 (6.4) | 0 | 75 (3.0) | 0 | <0.001 |
| Warfarin | 705 (6.8) | 0 | 270 (10.7) | 0 | <0.001 |
| DOACs | 615 (6.0) | 0 | 131 (5.2) | 0 | 0.140 |
| Diagnosis |  |  |  |  |  |
| CT of the abdomen/pelvis | 7,149 (69.1) | 0 | 507 (20.1) | NA | <0.001 |
| Contrast enhanced CT | 5,240 (73.3) | 0 | 149 (5.9) | NA | <0.001 |
| 1st colonoscopy | 9,064 (87.6) | 0 | 726 (29.3) | 47 (1.9) | <0.001 |
| 2nd colonoscopy | 1,992 (19.2) | 0 | 44 (1.7) | NA | <0.001 |
| Colonic diverticular bleeding | 6,575 (63.6) | 0 | 668 (26.4) | NA | <0.001 |
| Ischemic colitis | 941 (9.1) | 0 | 85 (3.4) | NA | <0.001 |
| Post-procedure bleeding | 463 (4.5) | 0 | 77 (3.0) | NA | 0.001 |
| Post-ESD bleeding | 140 (1.4) | 0 | 49 (1.9) | NA | 0.028 |
| Rectal ulcer | 257 (2.5) | 0 | 62 (2.5) | NA | 0.925 |
| IBD | 210 (2.0) | 0 | 305 (12.1) | NA | <0.001 |
| Hemorrhoid bleeding | 184 (1.8) | 0 | 25 (1.0) | NA | 0.005 |
| Other colorectal tumor | 9 (0.1) | 0 | 64 (2.5) | NA | <0.001 |
| Colorectal polyp | 37 (0.4) | 0 | 41 (1.6) | NA | <0.001 |
| Other diagnosis | 37 (0.4) | 0 | 576 (22.8) | NA | <0.001 |
| Unknown | 526 (5.1) | 0 | 67 (2.7) |  | <0.001 |
| Clinical outcomes |  |  |  |  |  |
| In-hospital rebleeding | 1,573 (15.2) | 0 | 343 (14.3) | 126 (5.0) | 0.251 |
| Acute coronary syndrome | 19 (0.2) | 0 | 7 (0.3) | 0 | 0.35 |
| Cerebrovascular disease | 28 (0.3) | 0 | 2 (0.1) | 0 | 0.073 |
| In-hospital death | 97 (0.9) | 0 | 85 (3.4) | 36 (1.4) | <0.001 |
| Blood transfusion | 3,080 (29.8) | 0 | 666 (26.7) | 35 (1.4) | 0.003 |
| Blood transfusion received ≥ 4 units | 2,291 (22.2) | 0 | 258 (10.3) | 35 (1.4) | <0.001 |
| Length of stay, median (IQR, range) | 7 (5-11, 0-229) | 0 | 3 (1–7) | 156 (6.2) | NA |

Abbreviations. IQR, interquartile range; LGIB, lower gastrointestinal bleeding; TIA, transient ischemic attack; COPD, chronic obstructive pulmonary disease; NSAIDs, non-steroidal anti-inflammatory drugs; DOACs, direct oral anticoagulants; IBD, inflammatory bowel disease; ESD, endoscopic submucosal dissection
